# Refining E-detectioN Tools for Emerging mental disoRders (ENTER) to identify young people at clinical high risk for psychosis in the community: model development and internal validation

**DOI:** 10.64898/2026.10.05.26364756

**Authors:** Dominic Oliver, Paolo Fusar-Poli, Gina Brunner, Kate Haining, Andrés Estradé, Matilda Azis, Bianca Bianciardi, Saskia Cooper, Soumita Ramesh, Phoebe Wallman, Xinyi Liang, Yanakan Logeswaran, Riccardo Stefanelli, Stefano Damiani, Antonio Melillo, Umberto Provenzani, Philip McGuire, Stephen Brewster, Shaun Macdonald, Patrizia Di Campli San Vito, Alex McConnachie, Caroline Haig, Thomas Spencer, Kelly Diederen, Peter J. Uhlhaas

## Abstract

Early detection of individuals at clinical high risk for psychosis (CHR-P) improves access to preventive care. We developed and internally validated the E-DetectioN Tool for Emerging mental disoRders (ENTER), a multimodal digital screener designed to improve community identification of CHR-P compared with an established approach (PQ-16 cut-off). Participants aged 12-35 years from the UK and Italy completed the online ENTER screener, assessing attenuated psychotic symptoms, processing speed and environmental risk factors. Participants above the PQ-16 cut-off and a random subset below were assessed with the Comprehensive Assessment of At Risk Mental States (CAARMS). PQ-16 cut-off and ENTER models were developed using (penalised) logistic regression, random forest and extreme gradient boosting (XGBoost) within a repeated nested cross-validation framework with semi-supervised learning incorporating unlabelled data. Models were evaluated by discrimination, calibration and clinical utility. 2,522 participants completed the ENTER screener. 495 underwent CAARMS assessments. 124 (25.1%) met CHR-P criteria; 2,027 participants were not assessed with the CAARMS (unlabelled). ENTER showed improved discrimination (C=0.744, 95%CI: 0.697-0.790) compared to the PQ-16 cut-off (C=0.625, 95%CI: 0.570-0.682, p=0.003) and similar calibration (ENTER: slope=1.02, 95%CI: 0.78-1.32; PQ-16 cut-off: slope=0.39, 95%CI: 0.19-0.52, p=0.074). At a clinically relevant threshold, ENTER identified approximately 2 additional CHR-P individuals per 100 assessments compared with the PQ-16 cut-off. Compared with established PQ-16 cut-off pre-screening, ENTER showed higher performance for community identification of CHR-P individuals. Multimodal digital screening might support more informative community triage before specialist assessment. External validation and implementation studies are required to establish real-world clinical utility.

## INTRODUCTION

Psychosis is one of the world’s leading causes of health-related disability,^1^ resulting in a high personal burden to affected individuals, their families and carers.^2^ The first episode of psychosis (FEP) typically occurs during adolescence or early adulthood,^3^ and is often preceded by a detectable clinical high risk for psychosis (CHR-P) phase,^4,5^ primarily characterised by attenuated psychotic symptoms.^6^ The CHR-P state is associated with an increased risk of developing psychosis, which reaches 25% at 3 years following the initial clinical assessment and continues to gradually increase over time to a cumulative risk of 46.2% at 18 years of follow-up,^7,8^ offering a unique opportunity for prevention. Identifying people early during the CHR-P phase is associated with improved long-term outcomes in patients with psychosis.^9,10^

However, effective detection of young people who might benefit from preventive care is a rate-limiting step for improving outcomes,^11^ with only 5-14% of first episode psychosis detected during the CHR-P phase.^12,13^ Currently, detection of CHR-P individuals is idiosyncratic, dependent on available resources of services.^14^ More systematic approaches to detection are critical to increase access to preventive care while ensuring psychosis risk within those detected remains high.^15^

Young people in the community may may experience subtle mental health problems and be seeking help online, representing a key target for more systematic digital detection strategies. Previous research from the Youth Mental Health Risk and Resilience Study (YouR-Study) has shown the potential of using the Prodromal Questionnaire (PQ-16),^16^ a 16-item self-report questionnaire about attenuated psychotic symptoms, online as a pre-screening tool to detect CHR-P individuals in the community with an acceptable area-under-the-curve of 0.74, driven by excellent sensitivity (0.81) and poor specificity (0.44) using a cut-off of ≥6.^17^ CHR-P individuals detected through this route had a 12.1% transition rate over three years.^18^ This cut-off is now typically used for pre-screening in CHR-P services.

Using data beyond symptomatology alone may improve performance. Distress associated with attenuated psychotic symptoms and impaired processing speed are linked to poorer clinical outcomes in CHR-P individuals.^19^ Similarly, CHR-P individuals accumulate various environmental, genetic and individual risk factors, which appear to modulate the neurobiological processes that drive psychosis aetiopathology.^20–22^ Measurement of these factors may therefore capture liability that is complementary to current symptom measures. We therefore incorporated symptom-related distress (PQ-16 distress), processing speed measured with the Digit Symbol Substitution Test (DSST),^23^ and environmental risk factors captured by the Psychosis Polyrisk Score (PPS).^24–26^

In this study, we developed and internally validated a clinical prediction model to improve community detection of CHR-P individuals using data from a self-administered online screener (E-DetectioN Tool for Emerging mental disoRders; ENTER), compared with the PQ-16 cut-off pre-screening approach as a benchmark.

## METHODS

### Participants

Participants aged 12-35 years were recruited to the ENTER study from Glasgow (Scotland), London (England), and Pavia (Italy) through social media, community advertising, universities and colleges. Interested individuals accessed study information online and completed electronic consent before undertaking the ENTER screener. The ENTER platform and promotional materials were informed by feedback from young experts by experience through the Young Person’s Mental Health Advisory Group at King’s College London. Recruitment started in April 2022 and ended in December 2024. The full recruitment strategy has been detailed in a previous publication.^27^

### Study design

#### Online screener

Participants were asked to complete the PQ-16^16^, DSST^23^ and PPS assessment.^24^

#### Assessment phase

Participants who met online inclusion criteria (PQ-16≥6) were invited through an email to participate in the second part of the study to determine CHR-P status using the Comprehensive Assessment of At Risk Mental States (CAARMS)^28^ 2016 version. A subset of participants who scored below the PQ-16 threshold were also randomly selected to be assessed with the CAARMS.

#### Ethics

Ethical approval was gained from East Midlands – Leicester Central Research Ethics Committee (UK sites; 21/EM/0283) and the Ethical Committee of Istituto di Ricovero e Cura a Carattere Scientifico San Matteo (Pavia; 0044135/22). All participants gave written informed consent. The study was registered on ClinicalTrials.gov (NCT05127278).

#### Statistical analysis

Model development and internal validation followed gold standard methodological guidelines^29^ and the Transparent Reporting of a Multivariable Prediction Model for Individual Prognosis or Diagnosis (TRIPOD+AI; eTable 1).^30^ All analyses were conducted in R version 4.4.2.

#### Model development

We aimed to develop models to discriminate between individuals who met CHR-P or psychosis criteria (CHR-P+) based on a gold-standard clinical interview (CAARMS) vs. those who did not (CHR-P-). As a first step, we used the PQ-16 cut-off typically used for pre-screening (≥6 symptoms endorsed). As a second step, we developed the ENTER multimodal screener with *a priori*-selected predictors: PQ-16 score, PQ-16 distress score, DSST, and PPS predictors (20 environmental risk factors). Detailed predictor information is presented in eMethods 1 and eTable 2.

Sample size calculations^31,32^ were performed post hoc so accurate parameters were available for robust estimates.

#### Internal validation

Internal validation was performed using repeated nested cross-validation with 5 inner and outer folds and 10 inner and outer repetitions, as implemented in caret (version 6.0-93). In each outer repetition, labelled data were split into five folds. Within each outer iteration, four folds were used as the training set with the remainder retained as an independent labelled test set. Hyperparameter tuning and missing data imputation were performed within the cross-validation framework (separately for labelled training, labelled test and unlabelled training data) (eMethods 2).

As 80.4% of participants who completed the website screener did not participate in the assessment phase (i.e. were not interviewed) and the assessment phase sample was enriched for individuals above the PQ-16 threshold, we used a previously tested semi-supervised learning (SSL) framework^33^ (Figure 1) to incorporate unlabelled data (i.e. data from participants who completed the online screener but did not attend an assessment). Within each outer training set, an initial pseudolabelling model was fitted using labelled data only (i.e. where CHR-P status was known). This model was then applied to the unlabelled observations to generate risk estimates. Unlabelled observations in the highest and lowest 5% of risk estimates were respectively assigned pseudolabels of CHR-P+ and CHR-P-. Negative pseudolabels were additionally required to have a predicted probability of 0.30 or lower. This process was then iteratively repeated to the combination of labelled and pseudeolabelled data up to 20 times or until the sum of the weight of pseudolabels (sum of predicted probabilities) in the sample exceeded three times the labelled data. More detail on the SSL procedure is presented in eMethods 3.

**Figure 1.**
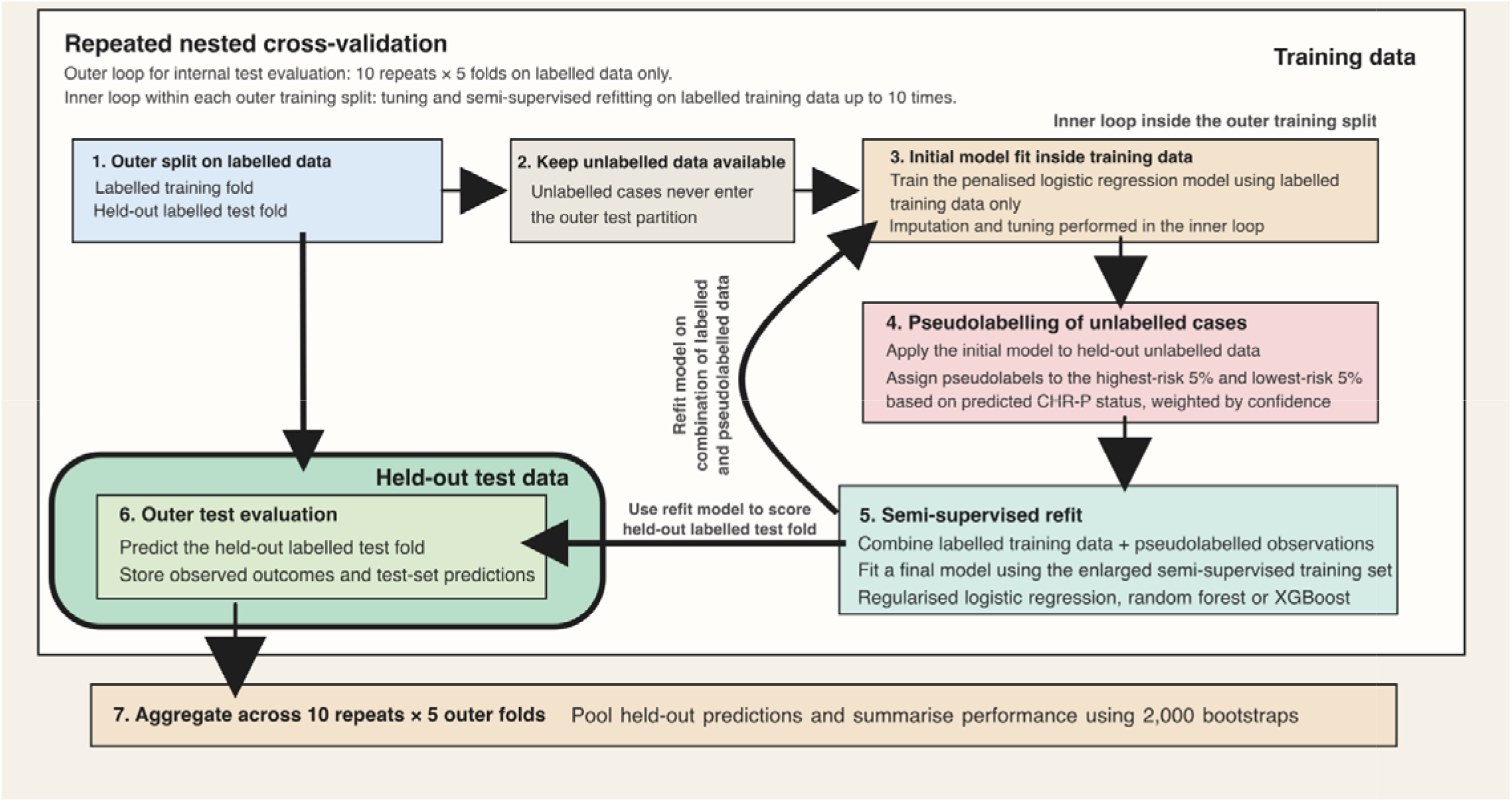
Semi-supervised learning framework implemented within repeated nested cross-validation. Within repeated nested cross-validation, an initial model trained on labelled data generated high-confidence pseudolabels for unlabelled observations. Pseudolabelled and labelled data were iteratively combined to create an expanded training set, which was used to develop final regularised logistic regression, random forest, and XGBoost models. Performance was evaluated on held-out labelled test folds only, with overall estimates derived from repeated cross-validation and 2,000 bootstrap resamples.

After the SSL procedure was complete, we used the final augmented outer training set (labelled plus weighted pseudolabelled data) to fit a final model: (i) elastic net-penalised logistic regression (single predictor models did not include penalisation), (ii) random forest and (iii) extreme gradient boosting (XGBoost). The performance of these models was then tested on held-out labelled test data.

#### Model performance

Model performance was primarily assessed through discrimination and calibration. Discrimination reflects the ability of a model to differentiate between CHR-P+ and CHR-P– and was quantified using Harrell’s C-index,^17^ defined as the probability that the predicted risk of the outcome is higher for the case than for the control in a randomly selected case-control pair.^25^ Calibration reflects the agreement between predicted probabilities and observed outcome frequencies. This was summarised using the calibration intercept, which assesses whether predictions are systematically too high or too low on average (ideal value 0), and the calibration slope, which assesses whether predicted risks are too extreme or too modest across the risk spectrum (ideal value 1). We also recorded the Brier score as an overall measure of performance (ideal value 0).

We additionally calculated balanced accuracy, sensitivity, specificity, positive predictive value (PPV) and negative predictive value (NPV) to assess classification performance (i.e. describing the relationship between true positives, false positives, false negatives and true negatives given a risk estimate cut-off). To assess this, we selected the threshold that produced the optimal balanced accuracy and presented true and false positive rates across risk thresholds in a classification plot.

Participant-level risk estimates were aggregated to calculate means and 95%CIs for all performance metrics using 2,000 bootstraps. Direct comparisons were conducted between models used the same participant-level bootstrap samples, with 95%CIs and two-sided p-values calculated for the paired difference in each performance metric.

#### Assessment of clinical utility

We used decision curve analysis to quantify the net benefit of using each clinical prediction model. This indexes the difference between the clinical benefit of accurate predictions (e.g. detecting psychosis cases early) and the clinical harm of inaccurate predictions (e.g. providing unnecessary preventive care). As different clinical scenarios may correspond to different preferences regarding missing psychosis cases and providing unnecessary interventions, we plotted net benefit across a range of threshold probabilities (0-50%). Values closer to 0% represent greater concern for missing potential psychosis cases and values closer to 50% represent greater concern for providing unnecessary preventive care. The net benefits of using the model were plotted compared to default strategies of assessing all or no individuals at risk.^34^ Standardised net benefit was calculated as the net benefit divided by the incidence of the outcome, showing the percentage increase in net benefit at a given risk threshold. As the prevalence of CHR-P individuals is dependent on the underlying population they are sampled from, we tested the net benefit using the estimated prevalence of CHR-P individuals in our sample, extrapolating the performance metrics of the PQ-16 reported in the YouR study^17^ to all participants completing the ENTER screener (28.3%).

#### Coefficients and variable importance

Model coefficients were derived from logistic regression models and variable importance from random forest and XGBoost. More detail is presented in eMethods 4.

#### Sensitivity analyses

Subgroup analyses were conducted for the full models, estimating discrimination and calibration performance using risk estimates from held-out test data, stratified by age, country, gender and ethnicity.

To assess the utility of each modality, we fitted separate models for clinical (PQ-16 score and distress), cognitive (DSST) and environmental (PPS) predictors.

## RESULTS

2,522 participants completed the online screener and met quality control criteria, excluding participants considered to be potentially fraudulent or inauthentic, as well as lower-quality responses, such as those failing attention checks or not meeting time thresholds (see ^27^ for more detail). The mean age was 23.9 years (SD 4.7). 574 (22.8%) of the dataset were male, 1,805 (71.6%) were female, and 100 (4.0%) were non-binary and 15 (0.6%) identified as other gender (28 [1.1%] participants were missing gender data). For self-assigned ethnicity, the dataset included 1,762 (69.9%) White, 480 (19.0%) Asian, 113 (4.5%) Mixed, 92 (3.6%) Other and 70 (2.8%) Black participants (5 [0.2%] were missing ethnicity data). Sociodemographics for the sample are presented in Table 1.

**Table 1.**
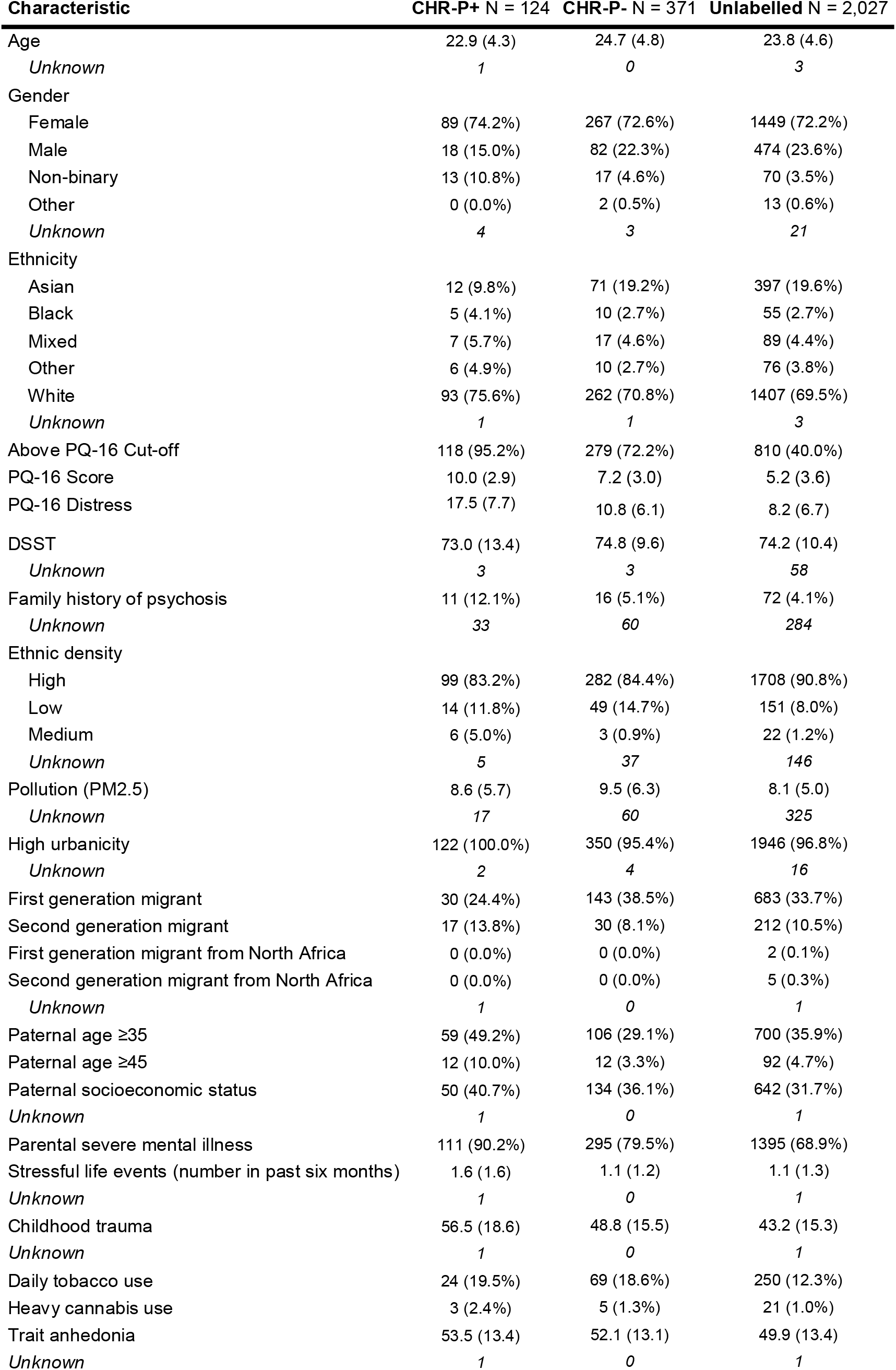
Sociodemographics table. Values are either presented as mean (SD) or n (%).

| Characteristic | **CHR-P+** N = 124 | **CHR-P-** N = 371 | **Unlabelled** N = 2,027 |
|---|---|---|---|
| Age | 22.9 (4.3) | 24.7 (4.8) | 23.8 (4.6) |
| *Unknown* | *1* | *0* | *3* |
| Gender |  |  |  |
| Female | 89 (74.2%) | 267 (72.6%) | 1449 (72.2%) |
| Male | 18 (15.0%) | 82 (22.3%) | 474 (23.6%) |
| Non-binary | 13 (10.8%) | 17 (4.6%) | 70 (3.5%) |
| Other | 0 (0.0%) | 2 (0.5%) | 13 (0.6%) |
| *Unknown* | *4* | *3* | *21* |
| Ethnicity |  |  |  |
| Asian | 12 (9.8%) | 71 (19.2%) | 397 (19.6%) |
| Black | 5 (4.1%) | 10 (2.7%) | 55 (2.7%) |
| Mixed | 7 (5.7%) | 17 (4.6%) | 89 (4.4%) |
| Other | 6 (4.9%) | 10 (2.7%) | 76 (3.8%) |
| White | 93 (75.6%) | 262 (70.8%) | 1407 (69.5%) |
| *Unknown* | *1* | *1* | *3* |
| Above PQ-16 Cut-off | 118 (95.2%) | 279 (72.2%) | 810 (40.0%) |
| PQ-16 Score | 10.0 (2.9) | 7.2 (3.0) | 5.2 (3.6) |
| PQ-16 Distress | 17.5 (7.7) | 10.8 (6.1) | 8.2 (6.7) |
| DSST | 73.0 (13.4) | 74.8 (9.6) | 74.2 (10.4) |
| *Unknown* | *3* | *3* | *58* |
| Family history of psychosis | 11 (12.1%) | 16 (5.1%) | 72 (4.1%) |
| *Unknown* | *33* | *60* | *284* |
| Ethnic density |  |  |  |
| High | 99 (83.2%) | 282 (84.4%) | 1708 (90.8%) |
| Low | 14 (11.8%) | 49 (14.7%) | 151 (8.0%) |
| Medium | 6 (5.0%) | 3 (0.9%) | 22 (1.2%) |
| *Unknown* | *5* | *37* | *146* |
| Pollution (PM2.5) | 8.6 (5.7) | 9.5 (6.3) | 8.1 (5.0) |
| *Unknown* | *17* | *60* | *325* |
| High urbanicity | 122 (100.0%) | 350 (95.4%) | 1946 (96.8%) |
| *Unknown* | *2* | *4* | *16* |
| First generation migrant | 30 (24.4%) | 143 (38.5%) | 683 (33.7%) |
| Second generation migrant | 17 (13.8%) | 30 (8.1%) | 212 (10.5%) |
| First generation migrant from North Africa | 0 (0.0%) | 0 (0.0%) | 2 (0.1%) |
| Second generation migrant from North Africa | 0 (0.0%) | 0 (0.0%) | 5 (0.3%) |
| *Unknown* | *1* | *0* | *1* |
| Paternal age ≥35 | 59 (49.2%) | 106 (29.1%) | 700 (35.9%) |
| Paternal age ≥45 | 12 (10.0%) | 12 (3.3%) | 92 (4.7%) |
| Paternal socioeconomic status | 50 (40.7%) | 134 (36.1%) | 642 (31.7%) |
| *Unknown* | *1* | *0* | *1* |
| Parental severe mental illness | 111 (90.2%) | 295 (79.5%) | 1395 (68.9%) |
| Stressful life events (number in past six months) | 1.6 (1.6) | 1.1 (1.2) | 1.1 (1.3) |
| *Unknown* | *1* | *0* | *1* |
| Childhood trauma | 56.5 (18.6) | 48.8 (15.5) | 43.2 (15.3) |
| *Unknown* | *1* | *0* | *1* |
| Daily tobacco use | 24 (19.5%) | 69 (18.6%) | 250 (12.3%) |
| Heavy cannabis use | 3 (2.4%) | 5 (1.3%) | 21 (1.0%) |
| Trait anhedonia | 53.5 (13.4) | 52.1 (13.1) | 49.9 (13.4) |
| *Unknown* | *1* | *0* | *1* |

1,207 participants (47.9%) met the published cut-off on the PQ-16 for psychosis risk with 397 attending an interview; a further 98 participants below the PQ-16 cut-off also attended. Of those who attended interviews, n=124 participants met CHR-P or psychosis criteria (CHR-P+) and n=371 did not (CHR-P-). 2,027 participants did not attend interviews (unlabelled). A study flow diagram is presented in eFigure 1.

With the prespecified common SSL rule, a mean of 2,027 (810 CHR-P+; mean 1.0 (SD=0.0) iteration) pseudolabelled participants were added for the PQ-16 cut-off model and 1,790 (895 CHR-P+; mean 20.0 (SD=0.0) iterations) for the ENTER multimodal screener.

The PQ-16 cut-off logistic regression model showed modest discrimination (C-index=0.625, 95%CI: 0.570-0.682) and poor calibration, with a calibration intercept of –1.31 (95%CI: –1.52--1.09), calibration slope of 0.39 (95%CI: 0.19-0.52), and Brier score of 0.27 (95%CI: 0.25-0.28) (Table 2, Figure 2).

**Figure 2.**
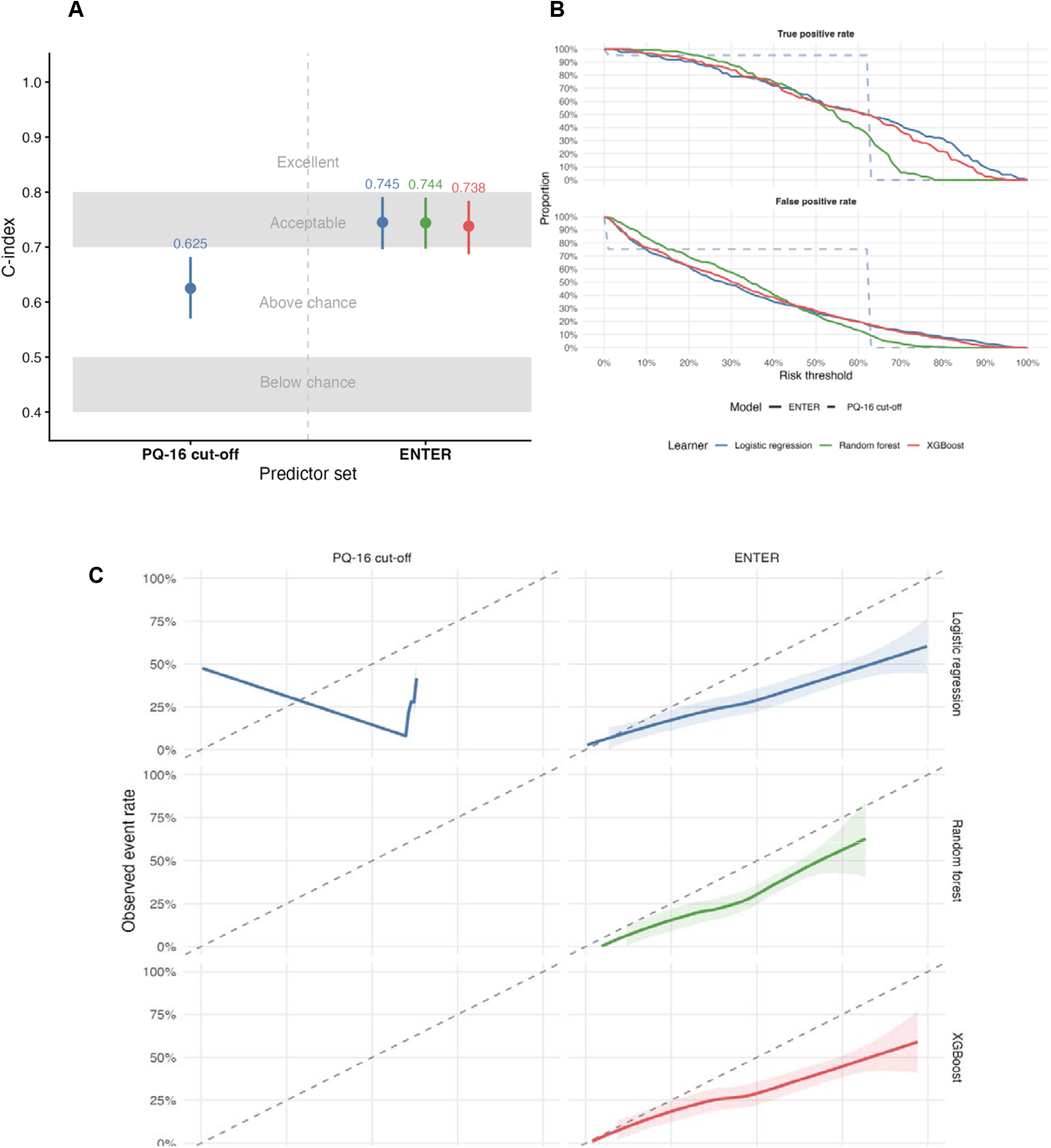
Performance summary of PQ-16 cutoff and ENTER multimodal models. **A** Discrimination performance. **B** Classification plot showcasing rates of true and false positives across a range of risk thresholds. **C** Calibration plots. Loess curves using pooled predictions and observed outcomes from held-out test data across cross-validation folds. Perfect calibration falls along the dashed diagonal line.

**Figure 3.**
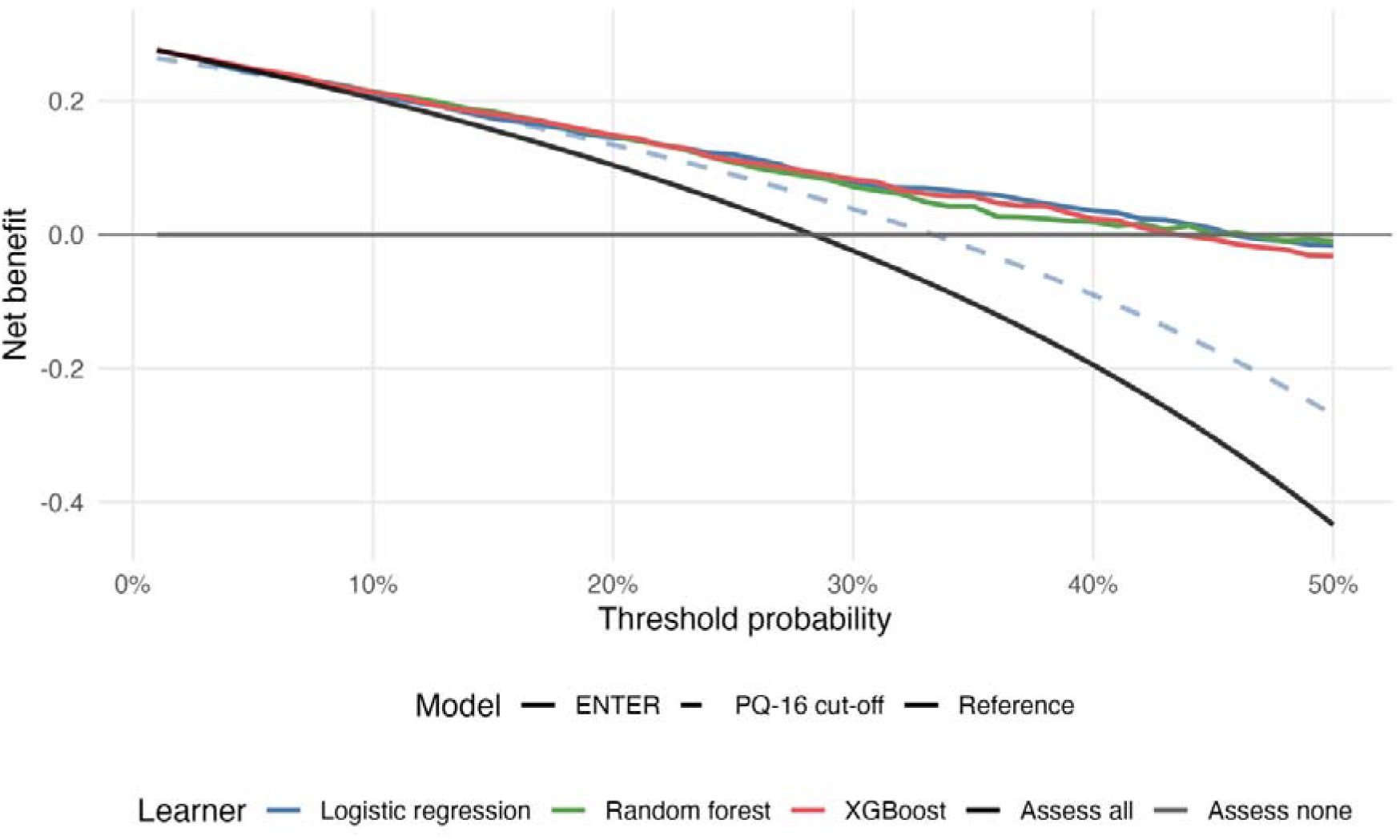
Decision curve analysis for PQ-16 cut-off (dashed lines) and ENTER (solid lines) models. The plots report net benefit (y axis) of the full prediction model across a range of risk thresholds (x axis) compared with assessing all individuals or in none assuming a prevalence of 28.3% within a help-seeking population.

**Table 2.** Model performance.

| Model | Model Type | C-index | Balanced Accuracy | Sensitivity | Specificity | PPV | NPV | Brier Score | Calibration Intercept | Calibration Slope |
|---|---|---|---|---|---|---|---|---|---|---|
| PQ-16 Cut-off | Logistic Regression | 0.625<br>(0.570-0.682) | 60.0%<br>(56.9%-62.8%) | 95.2%<br>(90.9%-98.4%) | 24.7%<br>(20.4%-29.4%) | 29.7%<br>(25.4%-34.3%) | 93.9%<br>(88.4%-98.0%) | 0.27<br>(0.25-0.28) | -1.31<br>(-1.52--1.09) | 0.39<br>(0.19-0.52) |
| ENTER | Logistic Regression | 0.745<br>(0.696-0.791) | 74.5%<br>(70.2%-78.5%) | 79.9%<br>(72.7%-86.6%) | 69.1%<br>(64.3%-73.7%) | 46.3%<br>(39.5%-52.8%) | 91.1%<br>(87.4%-94.3%) | 0.20<br>(0.18-0.22) | -1.04<br>(-1.33--0.76) | 0.55<br>(0.41-0.70) |
| ENTER | Random Forest | 0.744<br>(0.697-0.790) | 71.9%<br>(67.6%-76.3%) | 82.3%<br>(75.4%-88.9%) | 61.5%<br>(56.5%-66.6%) | 41.8%<br>(35.6%-48.0%) | 91.2%<br>(87.7%-94.7%) | 0.18<br>(0.17-0.20) | -0.75<br>(-0.97--0.54) | 1.02<br>(0.78-1.32) |
| ENTER | XGBoost | 0.738<br>(0.687-0.784) | 72.4%<br>(68.1%-76.5%) | 79.9%<br>(72.7%-86.9%) | 64.9%<br>(59.9%-69.8%) | 43.2%<br>(36.7%-49.8%) | 90.7%<br>(86.9%-94.1%) | 0.20<br>(0.18-0.22) | -0.97<br>(-1.24--0.70) | 0.63<br>(0.47-0.80) |

The ENTER multimodal screener showed significantly better discrimination than the PQ-16 cut-off (p<0.004; eTable 3). C-indices were 0.745 (95%CI: 0.696-0.791) for logistic regression, 0.744 (95%CI: 0.697-0.790) for random forest and 0.738 (95%CI: 0.687-0.784) for XGBoost (Table 2, Figure 2). There were no significant differences in discrimination performance between ENTER models (p>0.05, eTable 3).

Calibration was improved for the ENTER random forest relative to the other ENTER models, with a calibration intercept of –0.75 (95%CI: –0.97--0.54) and slope of 1.02 (95%CI: 0.78-1.32) (p<0.001, eTable 3). However, calibration intercept and slope were not improved compared to the PQ-16 cut-off model (p>0.05, eTable 3).

The ENTER multimodal screener showed significantly better overall performance compared to the PQ-16 cut-off (p<0.001, eTable 3). The ENTER random forest had the best overall performance, evidenced by the lowest Brier score (0.18, 95%CI: 0.17-0.20), compared with 0.20 (95%CI: 0.18-0.22) for both logistic regression and XGBoost (p<0.001, eTable 3).

PQ-16 cut-off and ENTER multimodal models met the sample size requirements to support the number of included predictors (eTable 4).

### Decision curve analysis

ENTER showed clinically useful net benefit across risk thresholds. If a CHR-P assessment were considered necessary at a risk threshold higher than 25% (i.e. missed detection of a CHR-P individual early is three times as harmful as an unnecessary assessment), the random forest model provided a net benefit of 6.4% over the most beneficial default strategy (assessing all or none) and 1.8% over the PQ-16 cut-off (eTable 5). This is equivalent to identifying approximately six additional CHR-P individuals per 100 people assessed compared to default strategies and two per 100 people assessed compared to the PQ-16 cut-off without increasing unnecessary assessments.

### Coefficients and variable importance

Migration-, parental– and deprivation-related factors, in addition to PQ-16-derived factors contributed most strongly to risk prediction in the ENTER multimodal screener. Full model coefficients and variable importance are presented in eTable 6.

### Sensitivity analyses

Discrimination and calibration did not differ significantly according to age, gender, country or ethnicity. However, overall performance measured by Brier score was poorer in non-binary participants (p<0.047) and Asian participants in the random forest model (p=0.03) (eTable 7-8).

ENTER showed significantly better discrimination performance than PQ-16 score (except logistic regression), PQ-16 distress (except logistic regression), DSST and PPS models. ENTER models generally showed lower Brier scores than unimodal models, with ENTER random forest generally showing better calibration. No unimodal model had significantly better performance than ENTER models (eTable 9-10; eFigure 2).

## DISCUSSION

This study demonstrates the potential value of a scalable digital screening approach (ENTER) for identifying CHR-P individuals in the community. Combining clinical, cognitive and environmental information improved the discrimination, calibration and potential clinical utility compared with the established PQ-16 cut-off, addressing the limited detection impact of current preventive approaches.

ENTER outperformed the PQ-16 cut-off, reinforcing the need to derive a broader picture of psychosis risk beyond symptoms alone. PQ-16 distress was a key predictor across multimodal models, underlining the importance of considering the intensity and impact of symptoms as well as their presence, in line with semi-structured clinical instruments. The importance of broader environmental risk factors is clinically relevant: variables related to migration, parents and deprivation contributed strongly to prediction performance.^22^ These findings are consistent with extensive epidemiological evidence linking social adversity, marginalisation and environmental stressors to psychosis risk.^21,22^ This supports evidence suggesting that the PPS can be a useful CHR-P detection tool, can discriminate between CHR-P individuals and healthy controls in international case-control samples (C=0.78-0.90),^25,35^ although such studies are not representative of community detection use case (unlike in the present study).

Decision curve analyses showed that ENTER demonstrated substantial clinical utility in community samples seeking help online. Compared with the PQ-16 cut-off, ENTER identified approximately two additional CHR-P individuals per 100 people assessed, corresponding to approximately 50 additional CHR-P individuals in this sample. This substantially increases pre-test risk, a key determinant of underlying psychosis risk within CHR-P populations.^15^

While there were no demonstrable differences in discrimination performance between ENTER models, the random forest ENTER model showed superior calibration and overall performance compared to the penalised logistic regression and XGBoost models. This may suggest that non-linear interactions between clinical, cognitive and environmental variables are relevant and may not be adequately captured by conventional regression. The evident heterogeneity of CHR-P samples suggests that psychosis onset is not caused by attenuated psychotic symptoms alone but is instead the result of a complex network of interacting clinical, cognitive and environmental factors.^21,36^ Machine learning methods may therefore offer advantages when modelling the complex, multidimensional architecture of psychosis risk.

The choice of learner also has implementation implications. At risk thresholds below 50%, the random forest model produced more true positives but also more false positives than the other ENTER models. Given the absence of clear differences in clinical utility, implementation may require prioritising either fewer false negatives or fewer unnecessary assessments depending on the setting. Accepting more false positives may be reasonable in well-resourced services seeking to maximise early detection, but less feasible where assessment capacity is limited.

This has important implications for CHR-P services and general preventive approaches. Currently, CHR-P services largely depend on referrals from clinicians,^8^ which may miss many young people experiencing emerging psychotic symptoms who do not present to specialist care. Digital screening tools, like ENTER, in combination with self-referral pathways offer the possibility of reaching larger and potentially underserved populations at earlier stages of symptom development, particularly when CHR-P services solely provide care to young people who are more likely to engage with digital health tools than traditional mental health services.^37^ ENTER could reduce access barriers and support allocation of specialist resources, although its broader information requirements create greater participant burden than the PQ-16 alone.

ENTER can also be conceptualised as one component within a broader unified digital detection framework. For example, P-Risk is an electronic health record-based clinical prediction model developed^38^ and externally validated^39^ in primary care, displaying robust performance (C=0.79). Similarly, a secondary care-based transdiagnostic risk calculator has been developed^40^, extensively externally validated (C=0.68-0.79^41–44^) and implemented in real world clinical practice^45,46^ (summarised in ^47^). Refined versions of the risk calculator have since been tested showing improved performance (C=0.85-0.89).^48,49^ While feasibility of these approaches may be limited by resourcing in CHR-P services to provide assessment, there is preliminary evidence that large language models can be used to augment and automate parts of the assessment pipeline.^50^ Together, these strategies can improve our ability to detect CHR-P individuals across different healthcare settings (i.e. ENTER in the community, P-Risk in primary care, transdiagnostic risk calculator in secondary care)^11^ at scale.

In addition, given that many environmental risk factors associated with psychosis also increase vulnerability to other mental disorders,^51^ there is an opportunity to expand ENTER for more transdiagnostic risk states outside of psychosis.^52^ This is particularly relevant in youth mental health, where early clinical presentations are often heterogeneous and non-specific, preceding multiple possible disorder trajectories.^53–55^ Consequently, digital tools such as ENTER may be well positioned to identify broader transdiagnostic risk states given the shared characteristics and overlap across prodromal phases of severe mental disorders.^56–58^ Existing work has also demonstrated the potential for clinical prediction models to identify transdiagnostic risk for outcomes such as psychosis and bipolar disorder more effectively than approaches relying on individual diagnoses alone.^59^ Validated frameworks for bipolar risk states, including the Semi-structured Interview for Bipolar At-Risk States (SIBARS),^60,61^ may provide a basis for extending this approach beyond psychosis.

The study also has important methodological implications. Only 20% of participants who completed the screener also completed gold-standard CAARMS assessments, reflecting the challenge in linking comprehensive semi-structured interviews to large-scale online data collection. Relying exclusively on labelled data would have risked producing models that were less representative of the intended deployment setting because interview attendance was not random. This would not have been detectable within the current dataset but would have reduced likelihood of generalisability in future samples. The adapted semi-supervised learning framework^33^ enabled incorporation of unlabelled data while retaining evaluation on real labelled participants. Such methods may help support scalable model development where fully labelled datasets are unavailable.

Several limitations should be considered. First, participants who attended interviews differed systematically from those who did not as interview invitations were based on the outcome of the PQ-16 cut-off, introducing selection bias. We have tried to mitigate this through assessing a random subset of participants below the PQ-16 cut-off so that labelled participants were not solely above the PQ-16 cut-off and employing semi-supervised learning. However, a larger, more complete sample would have been preferable. Second, self-report data remain vulnerable to measurement error and response biases, particularly as risk for fraudulent responses ranges from 3%-94% in online studies.^62^ We employed extensive quality control and fraud checks to mitigate the impact of this.^27^ Third, while recruitment was focused in sociodemographically diverse regions, the sample was predominantly female and White, which may limit generalisability to more diverse populations.

Future research should validate these findings in low-middle income settings.^63^ Fourth, while discrimination performance was promising, there was a general pattern of over-estimation of risk, which may be exacerbated in external validation. Fifth, subgroup analyses by ethnicity and gender were constrained by substantial class imbalance and small numbers, particularly in under 18s, non-White ethnic groups and non-binary participants, limiting robust assessment of algorithmic fairness. Finally, we were not able to externally validate our findings. Nevertheless, the use of data from three international sites, repeated nested cross-validation and adherence to methodological guidance^30^ provide stronger support for generalisability than many single-site development studies.

Scalable and fully digitised, self-administered multimodal digital screening approaches such as ENTER, integrating broader clinical, cognitive and environmental risk factors may improve detection of CHR-P individuals in the community. External validation and implementation studies are now required to confirm real-world clinical utility.

## Acknowledgements

DO and PM are supported by the National Institute for Health and Care Research (NIHR) Oxford Health Biomedical Research Centre. PFP is supported by the NIHR Maudsley Biomedical Research Centre. The views expressed are those of the author(s) and not necessarily those of the NHS, the NIHR or the Department of Health. PW and YL are supported by the UK Medical Research Council (MR/W006820/1). PFP is supported by #NEXTGENERATIONEU (NGEU), funded by the Ministry of University and Research (MUR), National Recovery and Resilience Plan (NRRP), project MNESYS (PE0000006) – A Multiscale integrated approach to the study of the nervous system in health and disease (DN. 1553 11.10.2022). KD is supported by the Academy of Medical Sciences (SBF007\100058).

## Author contributions

DO, PFP and PJU conceptualised the study. PFP, SB, SM, AMc and PU acquired funding. PFP, KH, MA, SC, PDCSV and PU set up the online screener. PFP, KH, MA, SD, UP, CH, TS, KD and PU coordinated data collection. GB, KH, AE, MA, PW, XL, RS, SD, AMe, UP and CH collected the data. AE, PW and RS accessed and verified the data. DO ran the statistical analyses. DO, PFP and PU drafted the original version of the manuscript. All authors edited and approved the final version of the manuscript.

## Funding

This study was funded by the Wellcome Trust (215793/Z/19/Z) awarded to PFP and PJU. Additional funding for collection of follow-up data was from the Academy for Medical Sciences through the Springboard Award (SBF007\100058). The funder of the study had no role in study design, data collection, data analysis, data interpretation or writing of the report.

## Conflicts of interest

DO has received consultancy fees from Google DeepMind outside of the current study. PFP has received research funds or personal fees from Lundbeck, Angelini, Menarini, Sunovion, Boehringer Ingelheim, Mindstrong, Proxymm Science, outside the current study. All other authors declare no conflicts of interest.

## Data availability

Data are available from the authors upon reasonable request to.

## Code availability

All analysis code is publicly available on GitHub: https://github.com/dapoliver/ENTER

## Notes

### Clinical Protocols

https://clinicaltrials.gov/study/NCT05127278?viewType=Card&rank=1

### Author Declarations

In the UK, ethical approval was given by East Midlands – Leicester Central Research Ethics Committee (Reference: 21/EM/0283). In Italy, ethical approval was given by Ethical Committee of Istituto di Ricovero e Cura a Carattere Scientifico (IRCCS) San Matteo (Reference: 0044135/22). All participants gave written informed consent.

### Summary of Updates

Results have been corrected across main text and supplementary after a minor issue in the analytic script was identified and resolved. There is no change in interpretation of results.

