## Supplementary Material for "Refining E-detectioN Tools for Emerging mental disoRders (ENTER) to identify young people at clinical high risk for psychosis in the community: model development and internal validation"

**eTable 1** TRIPOD+AI Checklist

**eMethods 1** List of all included risk/protective factors in PPS

**eTable 2** List of included factors, along with their definitions and cut-offs

**eMethods 2** Hyperparameter tuning and imputation

**eMethods 3** Semi-supervised learning methods

**eMethods 4** Model coefficients and variable importance

**eFigure 1** Study flow diagram

**eTable 3** Performance metric differences between models

**eTable 4** Sample size calculations

**eTable 5** Decision curve analysis

**eTable 6** Model coefficients and variable importance

**eTable 7** Sensitivity analysis: subgroup analysis

**eTable 8** Performance metric differences between subgroups in ENTER models

**eTable 9** Model performance of unimodal models

**eTable 10** Performance metric differences between unimodal and ENTER models

**eFigure 2** Performance summary of unimodal models

**eReferences**

| **eTable 1** TRIPOD+AI Checklist | | | | |
| --- | --- | --- | --- | --- |
| **Section/Topic Item Development Checklist item**  **/ evaluation**^1^ | | | | **Reported on page** |
| **TITLE** | | | |  |
| *Title* | 1 | D;E | Identify the study as developing or evaluating the performance of a multivariable prediction model, the target population, and the outcome to be predicted | 1 |
| **ABSTRACT** | | | | |
| *Abstract* | 2 | D;E | See TRIPOD+AI for Abstracts checklist | 2 |
| **INTRODUCTION** | | | | |
| *Background* | 3a | D;E | Explain the healthcare context (including whether diagnostic or prognostic) and rationale for developing or evaluating the prediction model, including references to existing models | 4-5 |
|  | 3b | D;E | Describe the target population and the intended purpose of the prediction model in the context of the care pathway, including its intended users (e.g., healthcare professionals, patients, public) | 4 |
|  | 3c | D;E | Describe any known health inequalities between sociodemographic groups | N/A |
| *Objectives* | 4 | D;E | Specify the study objectives, including whether the study describes the development or validation of a prediction model (or both) | 5 |
| **METHODS** | | | | |
| *Data* | 5a | D;E | Describe the sources of data separately for the development and evaluation datasets (e.g., randomised trial, cohort, routine care or registry data), the rationale for using these data, and representativeness of the data | 5 |
|  | 5b | D;E | Specify the dates of the collected participant data, including start and end of participant accrual; and, if applicable, end of follow-up | 5 |
| *Participants* | 6a | D;E | Specify key elements of the study setting (e.g., primary care, secondary care, general population)  including the number and location of centres | 5 |
|  | 6b | D;E | Describe the eligibility criteria for study participants | 5 |
|  | 6c | D;E | Give details of any treatments received, and how they were handled during model development or evaluation, if relevant | N/A |
| *Data preparation* | 7 | D;E | Describe any data pre-processing and quality checking, including whether this was similar across  relevant sociodemographic groups | eMethods 1-2 |
| *Outcome* | 8a | D;E | Clearly define the outcome that is being predicted and the time horizon, including how and when assessed, the rationale for choosing this outcome, and whether the method of outcome assessment is consistent across sociodemographic groups | 6 |
|  | 8b | D;E | If outcome assessment requires subjective interpretation, describe the qualifications and demographic characteristics of the outcome assessors | N/A |
|  | 8c | D;E | Report any actions to blind assessment of the outcome to be predicted | N/A |
| *Predictors* | 9a | D | Describe the choice of initial predictors (e.g., literature, previous models, all available predictors) and  any pre-selection of predictors before model building | eMethods 1; eTable 1 |
|  | 9b | D;E | Clearly define all predictors, including how and when they were measured (and any actions to blind assessment of predictors for the outcome and other predictors) | 6; eMethods 1; eTable 1 |
|  | 9c | D;E | If predictor measurement requires subjective interpretation, describe the qualifications and demographic characteristics of the predictor assessors | N/A |
| *Sample size* | 10 | D;E | Explain how the study size was arrived at (separately for development and evaluation), and justify that  the study size was sufficient to answer the research question. Include details of any sample size calculation | 7 |
| *Missing data* | 11 | D;E | Describe how missing data were handled. Provide reasons for omitting any data | eMethods 2 |
| *Analytical methods* | 12a | D | Describe how the data were used (e.g., for development and evaluation of model performance) in the analysis, including whether the data were partitioned, considering any sample size requirements | 6-7; eMethod 2-3 |
|  | 12b | D | Depending on the type of model, describe how predictors were handled in the analyses (functional form, rescaling, transformation, or any standardisation). | N/A |
|  | 12c | D | Specify the type of model, rationale^2^, all model-building steps, including any hyperparameter tuning,  and method for internal validation | 6-7; eMethod 2-3 |
|  | 12d | D;E | Describe if and how any heterogeneity in estimates of model parameter values and model performance was handled and quantified across clusters (e.g., hospitals, countries). See TRIPOD-Cluster for additional considerations^3^ | N/A |
|  | 12e | D;E | Specify all measures and plots used (and their rationale) to evaluate model performance (e.g., discrimination, calibration, clinical utility) and, if relevant, to compare multiple models | 8-9 |
|  | 12f | E | Describe any model updating (e.g., recalibration) arising from the model evaluation, either overall or for particular sociodemographic groups or settings | N/A |
|  | 12g | E | For model evaluation, describe how the model predictions were calculated (e.g., formula, code, object, application programming interface) | 8 |
| *Class imbalance* | 13 | D;E | If class imbalance methods were used, state why and how this was done, and any subsequent methods to recalibrate the model or the model predictions | N/A |
| *Fairness* | 14 | D;E | Describe any approaches that were used to address model fairness and their rationale | 9 |
| *Model output* | 15 | D | Specify the output of the prediction model (e.g., probabilities, classification). Provide details and  rationale for any classification and how the thresholds were identified | 7 |

^1^ D=items relevant only to the development of a prediction model; E=items relating solely to the evaluation of a prediction model; D;E=items applicable to both the development and evaluation of a prediction model

^2^ Separately for all model building approaches.

^3^ TRIPOD-Cluster is a checklist of reporting recommendations for studies developing or validating models that explicitly account for clustering or explore heterogeneity in model performance (eg, at different hospitals or centres). Debray et al, BMJ 2023; 380: e071018 [DOI: 10.1136/bmj-2022-071018]

| *Training versus*  *evaluation* | 16 | D;E | Identify any differences between the development and evaluation data in healthcare setting, eligibility criteria, outcome, and predictors | N/A |
| --- | --- | --- | --- | --- |
| *Ethical approval* | 17 | D;E | Name the institutional research board or ethics committee that approved the study and describe the participant-informed consent or the ethics committee waiver of informed consent | 6 |
| **OPEN SCIENCE** | | | | |
| *Funding* | 18a | D;E | Give the source of funding and the role of the funders for the present study | 16 |
| *Conflicts of interest* | 18b | D;E | Declare any conflicts of interest and financial disclosures for all authors | 16 |
| *Protocol* | 18c | D;E | Indicate where the study protocol can be accessed or state that a protocol was not prepared | 6 |
| *Registration* | 18d | D;E | Provide registration information for the study, including register name and registration number, or state that the study was not registered | 6 |
| *Data sharing* | 18e | D;E | Provide details of the availability of the study data | N/A |
| *Code sharing* | 18f | D;E | Provide details of the availability of the analytical code^4^ | 16 |
| **PATIENT & PUBLIC INVOLVEMENT** | | | | |
| *Patient & Public Involvement* | 19 | D;E | Provide details of any patient and public involvement during the design, conduct, reporting, interpretation, or dissemination of the study or state no involvement. | 5 |
| **RESULTS** | | | | |
| *Participants* | 20a | D;E | Describe the flow of participants through the study, including the number of participants with and without the outcome and, if applicable, a summary of the follow-up time. A diagram may be helpful. | 9; eFigure 1 |
|  | 20b | D;E | Report the characteristics overall and, where applicable, for each data source or setting, including the key dates, key predictors (including demographics), treatments received, sample size, number of outcome events, follow-up time, and amount of missing data. A table may be helpful. Report any differences across key demographic groups. | Table 1 |
|  | 20c | E | For model evaluation, show a comparison with the development data of the distribution of important predictors (demographics, predictors, and outcome). | N/A |
| *Model development* | 21 | D;E | Specify the number of participants and outcome events in each analysis (e.g., for model development, hyperparameter tuning, model evaluation) | 10 |
| *Model specification* | 22 | D | Provide details of the full prediction model (e.g., formula, code, object, application programming interface) to allow predictions in new individuals and to enable third-party evaluation and implementation, including any restrictions to access or re-use (e.g., freely available, proprietary)^5^ | eTable 4 |
| *Model performance* | 23a | D;E | Report model performance estimates with confidence intervals, including for any key subgroups (e.g., sociodemographic). Consider plots to aid presentation. | 10-11; Figure 2-3; Table 2; eTable 3 |
|  | 23b | D;E | If examined, report results of any heterogeneity in model performance across clusters. See TRIPOD Cluster for additional details^3^. | N/A |
| *Model updating* | 24 | E | Report the results from any model updating, including the updated model and subsequent performance | N/A |
| **DISCUSSION** | | | | |
| *Interpretation* | 25 | D;E | Give an overall interpretation of the main results, including issues of fairness in the context of the  objectives and previous studies | 11-14 |
| *Limitations* | 26 | D;E | Discuss any limitations of the study (such as a non-representative sample, sample size, overfitting, missing data) and their effects on any biases, statistical uncertainty, and generalizability | 15 |
| *Usability of the model in the context of current care* | 27a | D | Describe how poor quality or unavailable input data (e.g., predictor values) should be assessed and handled when implementing the prediction model | N/A |
|  | 27b | D | Specify whether users will be required to interact in the handling of the input data or use of the model,  and what level of expertise is required of users | N/A |
|  | 27c | D;E | Discuss any next steps for future research, with a specific view to applicability and generalizability of the model | 13-15 |

From: Collins GS, Moons KGM, Dhiman P, et al. *BMJ* 2024;385:e078378. doi:10.1136/bmj-2023-078378

^4^ This relates to the analysis code, for example, any data cleaning, feature engineering, model building, evaluation.

^5^ This relates to the code to implement the model to get estimates of risk for a new individual.

**eMethods 1** List of all included risk/protective factors in PPS.

We considered 17 factors that met the highest hierarchy of evidence (i.e. class I–III) for association with psychotic disorders, as detailed in an umbrella review of risk and protective factors for psychosis^1^. We then excluded those factors (n=6) that could not easily be measured at scale due to cost (Toxoplasma Gondii IgG), extended assessment time (premorbid IQ) or limited reliability of self-report (olfactory identification ability, minor physical anomalies). We also excluded the CHR-P state to investigate the PPS independently from this construct. This left 12 remaining class I-III risk factors. In addition to these risk factors, we additionally included some class IV risk factors that either overlapped with included factors or could be recorded at low cost, high reliability and limited assessment time (n=11).

Simple adaptations were added to the model to account for interdependencies in exposures, such as the multiple risk/protective factors associated with immigration status. For example, individuals cannot be exposed to certain immigration-based risk/protective factors in conjunction with each other. Immigrants cannot be both first generation and second-generation, and North African immigrants have to be either first- or second-generation immigrants. We combined these factors following this logic and assuming that the proportion and extra risk of North African immigrants is similar in first- and second-generation immigrants^2^. Factors related to ethnicity have similar logical dependencies between them, e.g. Black Caribbean is a non-White ethnicity, and individuals cannot be from a low ethnic density area, from a medium density area and from a high ethnic density area at the same time. We combined these factors again following this logic and assumed that the proportion and extra risk of Black Caribbean individuals between non-White ethnicity individuals is similar in low, medium and high ethnic density areas. Limitations of the PPS were previously presented^3,4^.

Of the 21 previously defined PPS factors, data were not available for hearing problems in the past 12 months.

Data on ethnic density, pollution and urbanicity were generated through area-level data linked to participants’ home addresses.

We did not use the weightings associated with the original PPS factors, as the associated psychosis risk in CHR-P individuals likely differs to the generated equivalent Odds Ratios from the umbrella review^1^**.**

**Processing of PPS variables**

Raw PPS questionnaire data were processed using a standardised pipeline implemented separately for the UK and Italian datasets. Date variables were converted into a common format and questionnaire items requiring reverse scoring (e.g., Childhood Trauma Questionnaire and Temporal Experience of Pleasure Scale items) were recoded before score derivation. Duplicate records were removed to ensure one observation per participant. Demographic and clinical risk factors were then transformed into harmonised variables corresponding to the PPS framework, including age, gender, ethnicity, migrant status, paternal age, paternal socioeconomic status, parental severe mental illness, adult life events, tobacco use, cannabis use, childhood trauma and trait anhedonia. Binary indicators were derived using predefined questionnaire thresholds, while continuous measures (e.g., childhood trauma and anhedonia) were calculated as summed scores.

Environmental risk factors were linked to participants using postcode- or municipality-level geographical information. Ethnic density, urbanicity, and air pollution exposure were derived from publicly available census and administrative datasets. In the UK dataset, participants were linked to small-area census geographies using postcode lookup tables, enabling derivation of ethnic density categories, population density measures, and PM2.5 exposure estimates based on the nearest available pollution monitoring grid. In the Italian dataset, equivalent variables were generated using municipality-level demographic, population density, and air pollution data linked through postcode information. Continuous environmental measures were categorised using predefined or distribution-based thresholds consistent with the original PPS methodology. Following derivation of all component variables, all processed variables were exported for subsequent modelling analyses.

***Class I***

1. Black‐Caribbean ethnicity in England (and Black African ethnicity in England – class IV)

***Class II***

2. Ethnic minority in low ethnic density area
3. Second generation immigrants
4. Trait anhedonia

***Class III***

5. Childhood trauma
6. Ethnic minority in high ethnic density area
7. First generation immigrants
8. Non‐right handedness
9. North African immigrants in Europe
10. Urbanicity

***Class IV***

11. Adult life events
12. Other ethnicity (Asian/Mixed/Other White ethnicity in England)
13. Heavy cannabis use
14. Low paternal socio‐economic status
15. Parental severe mental illness
16. Tobacco use
17. Pollution
18. Male & aged 25-35 years old

| **eTable 2** List of included factors, along with their definitions and cut-offs | | | |
| --- | --- | --- | --- |
| **Risk factor assessed (Classification of evidence; Meta-analytical reference)** | **Assessment tool; Reference** | **Scoring (Classification of evidence; meta-analytical reference)** | **Continuous/Categorical (Cutoff)** |
| Age/Gender^5^ (IV) |  |  |  |
| Non-right-handedness^6^ (III) |  |  |  |
| Pollution^7^ (IV) |  | Local census data | Continuous: PM2.5 exposure |
| Urbanicity^8^ (III) |  | Local census data | Categorical: 300 people per square kilometre (km^2^) or 3 people per hectare |
| Ethnic density |  | Local census data | Categorical:  Low: Lowest 33^rd^ percentile  Medium: Middle 33^rd^ percentile  High: Highest 33^rd^ percentile |
| Black Ethnicity^9^ (I) |  | In low ethnic density area^10^ (II) |  |
|  |  | In medium ethnic density area |  |
|  |  | In high ethnic density area^10^ (III) |  |
| Other Ethnicity^5^ (IV) |  | In low ethnic density area^10^ (II) |  |
|  |  | In medium ethnic density area |  |
|  |  | In high ethnic density area^10^ (III) |  |
| White |  |  |  |
| Not immigrant |  |  |  |
| 1st generation immigrant^11^ (III) |  | From North Africa^12^ (III) |  |
|  |  | From other regions |  |
| 2nd generation immigrant (II) |  | From North Africa^12^ (III) |  |
|  |  | From other regions |  |
| Paternal age <35 |  |  |  |
| Paternal age > 35^13^ (IV) |  |  |  |
| Paternal age > 45^13^ (IV) |  |  |  |
| Paternal SES^14^ (IV) |  |  |  |
| Parental Severe Mental Illness^15^ (IV) | Family Interview for Genetic Studies [FIGS]^16^ |  | Categorical: ≥1 disorder endorsed |
| Adult Life Events^17^ (IV) | Life Threatening Events Questionnaire [LTE-Q]^18^ |  | Categorical: Continuous |
| Tobacco use^19^ (IV) |  |  | Categorical: Daily smoker |
| Heavy cannabis use^20^ (IV) | Cannabis Experience Questionnaire |  | Categorical: More than once a week or every day |
| Childhood Trauma^21^ (III) | Childhood Trauma Questionnaire Short Form [CTQ-SF]^22^ | *Never true = 1, rarely true = 2, sometimes true = 3, often true = 4, Very often true =5. None = 5-40, Low = 41-55; Moderate = 56-72; Severe = 73+* | Continuous |
| Trait Anhedonia^23^ (II) | Temporal Experience of Pleasure Scale (TEPS)^24^ |  | Continuous |

**eMethods 2** Hyperparameter tuning and imputation of missing data

Hyperparameters were tuned using grid search within the inner folds of the cross-validation structure. For elastic-net penalised logistic regression, candidate alpha values were 0.25, 0.50, and 0.75, and candidate lambda values comprised 50 evenly spaced values from 0.001 to 0.1. Random forest models used 500 trees and tuned mtry (3, 6, 9, or 12), minimum node size (20, 50, 100, or 150 observations), and maximum tree depth (4, 6, or 8). XGBoost models tuned the number of boosting rounds (200, 400, or 800), maximum tree depth (2, 3, or 4), learning rate eta (0·01, 0·03, or 0·05), minimum loss reduction gamma (0, 1, or 5), subsample ratio (0·6, 0·8, or 0·9), column subsampling ratio per tree (0·4, 0·6, or 0·8), and minimum child weight (1, 2, or 5).

For elastic-net models, the configuration with the highest mean inner-fold C-index was selected. To reduce overfitting in the tree-based learners, we applied a one-standard-error rule for random forest and XGBoost: all configurations with mean C-index within one standard error of the highest-performing configuration were retained, and the most regularised configuration was selected for refitting on the complete outer training set before evaluation in the held-out outer fold.

Random forest imputation (missForest version 1.5; default parameters) was preferred with multiple imputation using chained equations (mice version 3.16.0; default parameters) if model did not converge, following investigation of missingness patterns using the naniar package (version 1.1.0).

**eMethods 3** Semi-supervised learning methods

Pseudolabelled observations were incorporated into model refitting with weights proportional to prediction confidence, defined as 2 x absolute(predicted probability - 0·5), while labelled observations retained unit weight. To prevent pseudolabelled data from dominating model estimation, the cumulative pseudolabel weight within each outer training set was capped at a prespecified multiple of the total labelled-data weight.

Because there was no objective ground truth for the optimal SSL augmentation rule, we conducted a tuning exercise using training folds only to identify a common semi-supervised augmentation rule that would not unduly favour either model.

Candidate values for the lower-tail probability gate for negative pseudolabels (0·30, 0·40, 0·50), the cumulative pseudolabel weight cap ratio (1·0, 1·5, 2·0, 2·5, 3·0 times the total labelled-data weight), and the proportion of unlabelled observations selected from each tail at each iteration (5%, 10%, 15%, 20%) were evaluated within the inner loop of nested cross-validation using labelled outer-training data and the associated unlabelled pool only. Pseudolabelling was performed using a LASSO-penalised logistic regression SSL learner, and the final prediction models were then refitted as penalised logistic regression, random forest or XGBoost models. On the basis of this tuning exercise, a common semi-supervised augmentation rule was selected and then fixed for the main analysis.

The final prespecified rule selected the upper and lower 5% of the unlabelled risk distribution at each iteration, used a negative pseudolabelling probability gate of 0·30, and capped cumulative pseudolabel weight at 3·0 times the total labelled-data weight. Tails were not class-balanced, and pseudolabel probabilities were not recalibrated before selection. This ensured that the held-out outer test fold remained uninvolved in both pseudolabelling-rule selection and model fitting.

**eMethods 4** Coefficients and variable importance

Because the primary semi-supervised learning procedure yielded multiple final models across the repeated resampling framework rather than a single fixed coefficient vector, a final model could not be obtained by fitting one model to the full dataset. To facilitate external validation, we therefore derived a pooled implementation model by aggregating the final coefficients from the repeated semi-supervised refits. For each predictor, coefficients were pooled on the linear predictor scale using the median as the primary summary, and coefficient variability across repeated fits was reported to quantify parameter stability. The pooled model was then applied to the held-out labelled test data and its performance was compared with that of the primary semi-supervised procedure.

For the penalised logistic regression models, predictor contribution was summarised using pooled coefficients derived across repeated semi-supervised refits. For the random forest and XGBoost models, predictor contribution was summarised using model-specific variable importance measures. Specifically, permutation importance was extracted from the random forest models, whereas gain, cover, and frequency were extracted from the XGBoost models. To quantify stability across repeated fits, these quantities were summarised across resampling iterations using their median, mean, standard deviation, and empirical 95% interval. These measures were interpreted as relative indicators of predictor contribution and ranking rather than as directly comparable effect sizes across modelling approaches.

**eFigure 1** Study flow diagram adapted from Wallman et al. (2025)^33^ showcasing participant flow in the UK (**A**) and Italy (**B**)

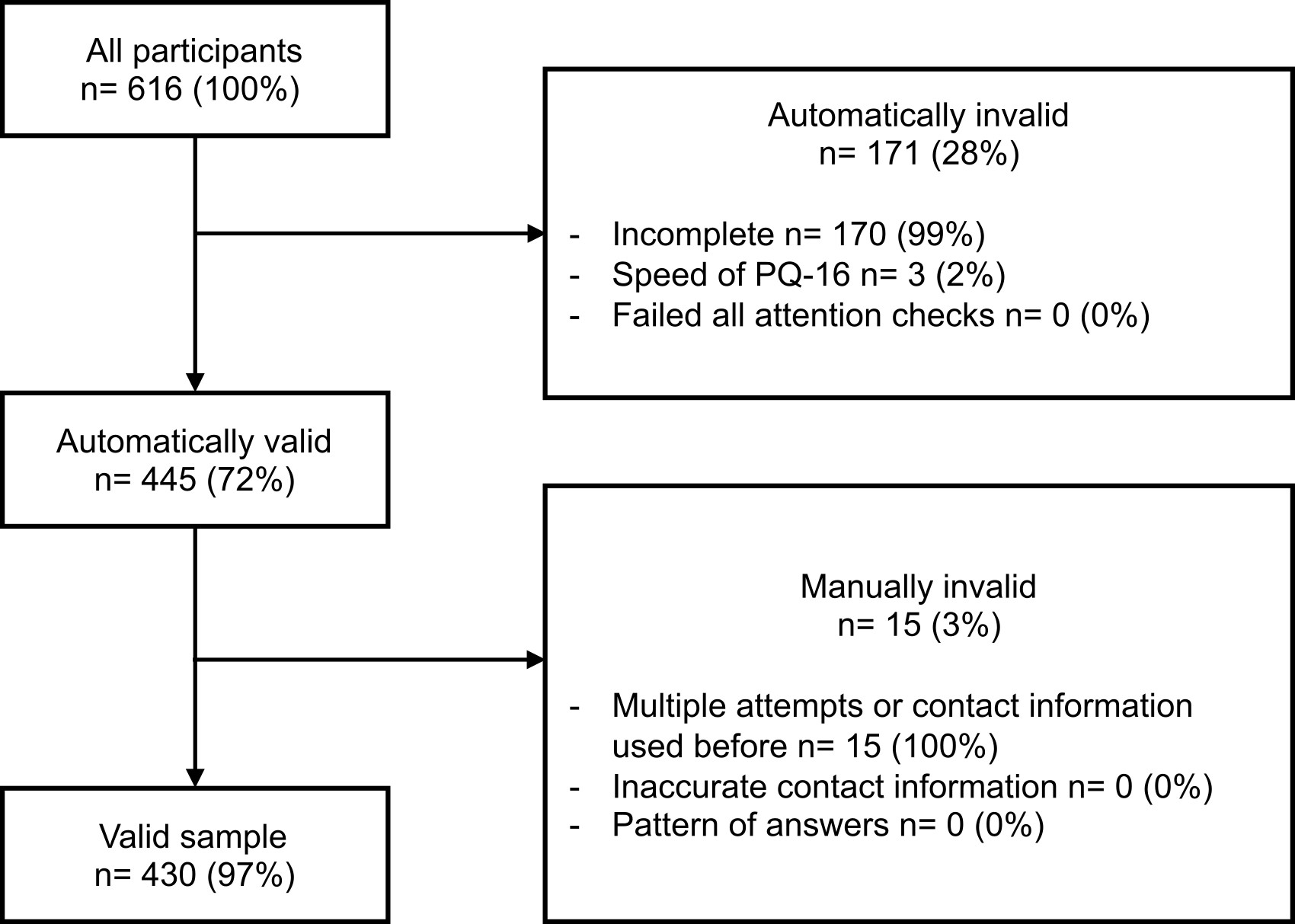

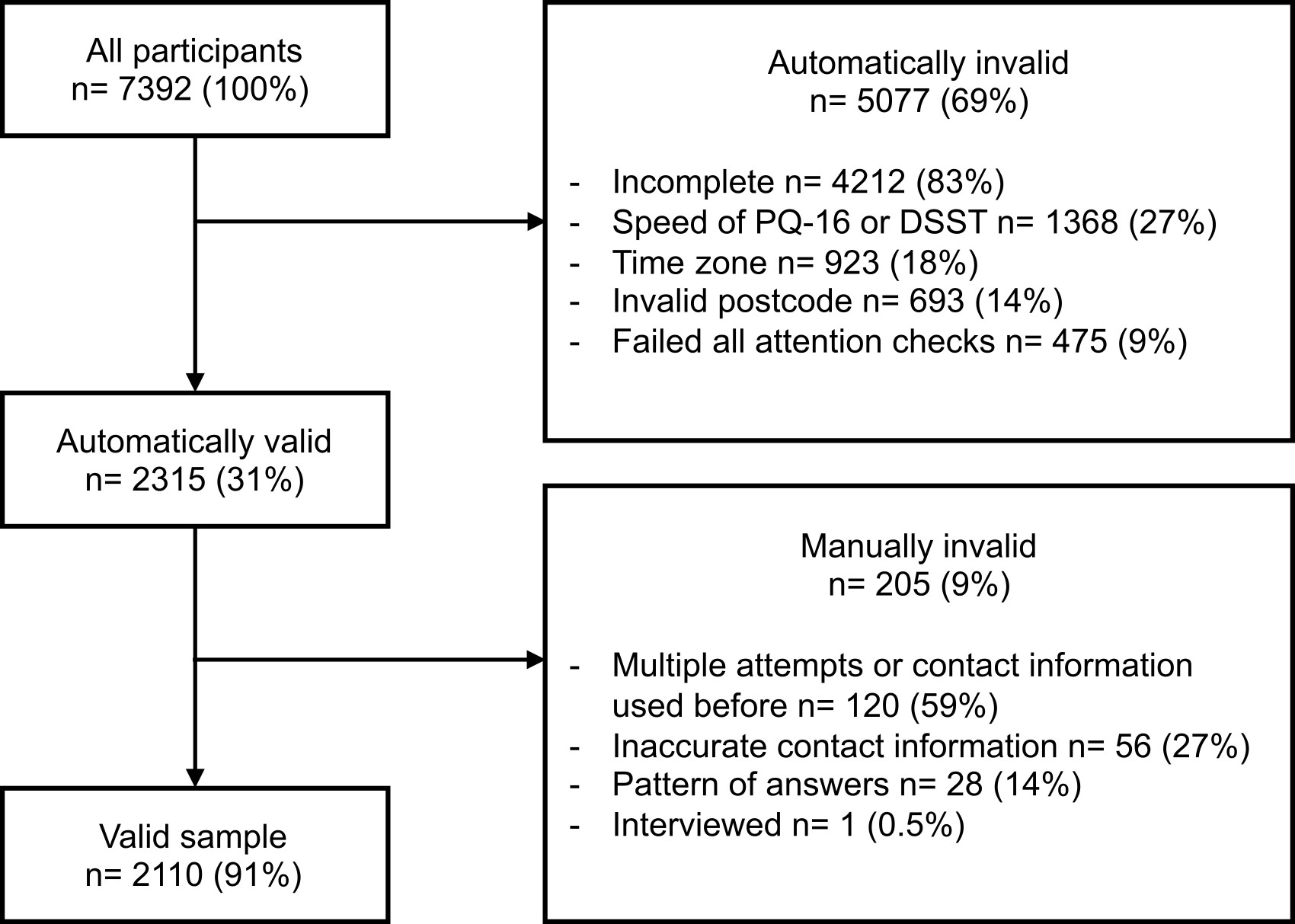

**B**

**A**

| **eTable 3** Bootstrapped performance metric differences between models. Statistically significant differences are highlighted in **bold.** | | | | | | |
| --- | --- | --- | --- | --- | --- | --- |
| **Model A** | **Model B** | **Metric** | **Difference** | **Lower 95%CI** | **Upper 95%CI** | **p-value** |
| **PQ-16 Cut-off Logistic Regression** | **ENTER Logistic Regression** | **C** | **-0.113** | **-0.179** | **-0.047** | **<0.001** |
| **PQ-16 Cut-off Logistic Regression** | **ENTER Logistic Regression** | **Balanced Accuracy** | **-0.127** | **-0.169** | **-0.084** | **<0.001** |
| **PQ-16 Cut-off Logistic Regression** | **ENTER Logistic Regression** | **Brier** | **0.062** | **0.039** | **0.084** | **<0.001** |
| PQ-16 Cut-off Logistic Regression | ENTER Logistic Regression | Calibration Intercept | -0.349 | -1.432 | 0.396 | 0.474 |
| PQ-16 Cut-off Logistic Regression | ENTER Logistic Regression | Calibration Slope | 0.151 | -1.161 | 1.973 | 0.951 |
| **PQ-16 Cut-off Logistic Regression** | **ENTER Random Forest** | **C** | **-0.108** | **-0.173** | **-0.042** | **0.003** |
| **PQ-16 Cut-off Logistic Regression** | **ENTER Random Forest** | **Balanced Accuracy** | **-0.129** | **-0.165** | **-0.093** | **<0.001** |
| **PQ-16 Cut-off Logistic Regression** | **ENTER Random Forest** | **Brier** | **0.080** | **0.063** | **0.098** | **<0.001** |
| **PQ-16 Cut-off Logistic Regression** | **ENTER Random Forest** | Calibration Intercept | 0.194 | -0.790 | 1.008 | 0.487 |
| **PQ-16 Cut-off Logistic Regression** | **ENTER Random Forest** | Calibration Slope | -1.393 | -2.914 | 0.312 | 0.082 |
| **PQ-16 Cut-off Logistic Regression** | **ENTER XGBoost** | **C** | **-0.103** | **-0.167** | **-0.035** | **0.003** |
| **PQ-16 Cut-off Logistic Regression** | **ENTER XGBoost** | **Balanced Accuracy** | **-0.112** | **-0.155** | **-0.069** | **<0.001** |
| **PQ-16 Cut-off Logistic Regression** | **ENTER XGBoost** | **Brier** | **0.062** | **0.041** | **0.083** | **<0.001** |
| PQ-16 Cut-off Logistic Regression | ENTER XGBoost | Calibration Intercept | -0.430 | -1.282 | 0.411 | 0.492 |
| PQ-16 Cut-off Logistic Regression | ENTER XGBoost | Calibration Slope | 0.228 | -1.239 | 1.711 | 0.961 |
| ENTER Logistic Regression | ENTER Random Forest | C | 0.005 | -0.016 | 0.025 | 0.644 |
| ENTER Logistic Regression | ENTER Random Forest | Balanced Accuracy | -0.003 | -0.039 | 0.032 | 0.878 |
| ENTER Logistic Regression | ENTER Random Forest | **Brier** | **0.018** | **0.007** | **0.030** | **0.002** |
| **ENTER Logistic Regression** | **ENTER Random Forest** | **Calibration Intercept** | **0.614** | **0.366** | **0.911** | **<0.001** |
| **ENTER Logistic Regression** | **ENTER Random Forest** | **Calibration Slope** | **-1.654** | **-2.313** | **-1.083** | **<0.001** |
| ENTER Logistic Regression | ENTER XGBoost | C | 0.011 | -0.002 | 0.024 | 0.107 |
| ENTER Logistic Regression | ENTER XGBoost | Balanced Accuracy | 0.015 | -0.019 | 0.050 | 0.401 |
| ENTER Logistic Regression | ENTER XGBoost | Brier | 0.000 | -0.006 | 0.006 | 0.985 |
| ENTER Logistic Regression | ENTER XGBoost | Calibration Intercept | 0.017 | -0.094 | 0.128 | 0.772 |
| ENTER Logistic Regression | ENTER XGBoost | Calibration Slope | -0.080 | -0.295 | 0.135 | 0.447 |
| ENTER Random Forest | ENTER XGBoost | C | 0.007 | -0.009 | 0.022 | 0.412 |
| ENTER Random Forest | ENTER XGBoost | Balanced Accuracy | 0.018 | -0.012 | 0.049 | 0.262 |
| ENTER Random Forest | ENTER XGBoost | **Brier** | **-0.019** | **-0.027** | **-0.010** | **<0.001** |
| **ENTER Random Forest** | **ENTER XGBoost** | **Calibration Intercept** | **-0.605** | **-0.864** | **-0.389** | **<0.001** |
| **ENTER Random Forest** | **ENTER XGBoost** | **Calibration Slope** | **1.591** | **1.103** | **2.159** | **<0.001** |

**eTable 4** Sample size calculations. Note observed n and events refer to the mean n and events following incorporation of pseudolabelled data.

| **Predictors** | **Model** | **N** | **Event rate** | **Predictors** | **Nagelkerke R^2^** | **Shrinkage** | **Criterion 1 n** | **Criterion 2 n** | **Criterion 3 n** | **Observed n minus required n** | **Observed events minus required events** |
| --- | --- | --- | --- | --- | --- | --- | --- | --- | --- | --- | --- |
| PQ-16 cut-off | Logistic regression | 2522 | 0.37 | 1 | 0.06 | 0.9 | 196 | 27 | 359 | 2163 | 801 |
| ENTER | Logistic regression | 2287 | 0.44 | 29 | 0.23 | 0.9 | 1393 | 694 | 380 | 894 | 396 |
| ENTER | Random forest | 2287 | 0.44 | 29 | 0.22 | 0.9 | 1457 | 697 | 380 | 830 | 368 |
| ENTER | XGBoost | 2287 | 0.44 | 29 | 0.21 | 0.9 | 1552 | 701 | 380 | 735 | 326 |

**eTable 5** Decision curve analysis

| Threshold | Predictors | Model | Net Benefit | Standardised Net Benefit |
| --- | --- | --- | --- | --- |
| 0.01 | PQ-16 Cut-off | Logistic Regression | -0.012 | -0.042 |
| 0.02 | PQ-16 Cut-off | Logistic Regression | -0.01 | -0.036 |
| 0.03 | PQ-16 Cut-off | Logistic Regression | -0.008 | -0.029 |
| 0.04 | PQ-16 Cut-off | Logistic Regression | -0.006 | -0.022 |
| 0.05 | PQ-16 Cut-off | Logistic Regression | -0.004 | -0.015 |
| 0.06 | PQ-16 Cut-off | Logistic Regression | -0.002 | -0.008 |
| 0.07 | PQ-16 Cut-off | Logistic Regression | 0 | -0.001 |
| 0.08 | PQ-16 Cut-off | Logistic Regression | 0.002 | 0.006 |
| 0.09 | PQ-16 Cut-off | Logistic Regression | 0.004 | 0.014 |
| 0.1 | PQ-16 Cut-off | Logistic Regression | 0.006 | 0.021 |
| 0.11 | PQ-16 Cut-off | Logistic Regression | 0.008 | 0.029 |
| 0.12 | PQ-16 Cut-off | Logistic Regression | 0.011 | 0.037 |
| 0.13 | PQ-16 Cut-off | Logistic Regression | 0.013 | 0.045 |
| 0.14 | PQ-16 Cut-off | Logistic Regression | 0.015 | 0.054 |
| 0.15 | PQ-16 Cut-off | Logistic Regression | 0.018 | 0.062 |
| 0.16 | PQ-16 Cut-off | Logistic Regression | 0.02 | 0.071 |
| 0.17 | PQ-16 Cut-off | Logistic Regression | 0.023 | 0.08 |
| 0.18 | PQ-16 Cut-off | Logistic Regression | 0.025 | 0.09 |
| 0.19 | PQ-16 Cut-off | Logistic Regression | 0.028 | 0.099 |
| 0.2 | PQ-16 Cut-off | Logistic Regression | 0.031 | 0.109 |
| 0.21 | PQ-16 Cut-off | Logistic Regression | 0.034 | 0.119 |
| 0.22 | PQ-16 Cut-off | Logistic Regression | 0.036 | 0.129 |
| 0.23 | PQ-16 Cut-off | Logistic Regression | 0.039 | 0.139 |
| 0.24 | PQ-16 Cut-off | Logistic Regression | 0.042 | 0.15 |
| 0.25 | PQ-16 Cut-off | Logistic Regression | 0.046 | 0.161 |
| 0.26 | PQ-16 Cut-off | Logistic Regression | 0.049 | 0.172 |
| 0.27 | PQ-16 Cut-off | Logistic Regression | 0.052 | 0.184 |
| 0.28 | PQ-16 Cut-off | Logistic Regression | 0.055 | 0.196 |
| 0.29 | PQ-16 Cut-off | Logistic Regression | 0.049 | 0.173 |
| 0.3 | PQ-16 Cut-off | Logistic Regression | 0.038 | 0.135 |
| 0.31 | PQ-16 Cut-off | Logistic Regression | 0.027 | 0.096 |
| 0.32 | PQ-16 Cut-off | Logistic Regression | 0.016 | 0.055 |
| 0.33 | PQ-16 Cut-off | Logistic Regression | 0.004 | 0.013 |
| 0.34 | PQ-16 Cut-off | Logistic Regression | -0.008 | -0.03 |
| 0.35 | PQ-16 Cut-off | Logistic Regression | -0.021 | -0.074 |
| 0.36 | PQ-16 Cut-off | Logistic Regression | -0.034 | -0.12 |
| 0.37 | PQ-16 Cut-off | Logistic Regression | -0.047 | -0.167 |
| 0.38 | PQ-16 Cut-off | Logistic Regression | -0.061 | -0.216 |
| 0.39 | PQ-16 Cut-off | Logistic Regression | -0.075 | -0.267 |
| 0.4 | PQ-16 Cut-off | Logistic Regression | -0.09 | -0.319 |
| 0.41 | PQ-16 Cut-off | Logistic Regression | -0.105 | -0.372 |
| 0.42 | PQ-16 Cut-off | Logistic Regression | -0.121 | -0.428 |
| 0.43 | PQ-16 Cut-off | Logistic Regression | -0.137 | -0.486 |
| 0.44 | PQ-16 Cut-off | Logistic Regression | -0.154 | -0.545 |
| 0.45 | PQ-16 Cut-off | Logistic Regression | -0.172 | -0.607 |
| 0.46 | PQ-16 Cut-off | Logistic Regression | -0.19 | -0.671 |
| 0.47 | PQ-16 Cut-off | Logistic Regression | -0.209 | -0.738 |
| 0.48 | PQ-16 Cut-off | Logistic Regression | -0.228 | -0.807 |
| 0.49 | PQ-16 Cut-off | Logistic Regression | -0.249 | -0.879 |
| 0.5 | PQ-16 Cut-off | Logistic Regression | -0.27 | -0.954 |
| 0 | ENTER | Logistic Regression | 0 | 0 |
| 0.01 | ENTER | Logistic Regression | 0 | 0 |
| 0.02 | ENTER | Logistic Regression | 0.001 | 0.002 |
| 0.03 | ENTER | Logistic Regression | -0.003 | -0.01 |
| 0.04 | ENTER | Logistic Regression | -0.003 | -0.012 |
| 0.05 | ENTER | Logistic Regression | -0.001 | -0.004 |
| 0.06 | ENTER | Logistic Regression | 0.001 | 0.005 |
| 0.07 | ENTER | Logistic Regression | 0.002 | 0.006 |
| 0.08 | ENTER | Logistic Regression | 0.005 | 0.016 |
| 0.09 | ENTER | Logistic Regression | 0.004 | 0.013 |
| 0.1 | ENTER | Logistic Regression | 0.007 | 0.024 |
| 0.11 | ENTER | Logistic Regression | 0.011 | 0.039 |
| 0.12 | ENTER | Logistic Regression | 0.012 | 0.042 |
| 0.13 | ENTER | Logistic Regression | 0.016 | 0.055 |
| 0.14 | ENTER | Logistic Regression | 0.016 | 0.057 |
| 0.15 | ENTER | Logistic Regression | 0.017 | 0.058 |
| 0.16 | ENTER | Logistic Regression | 0.022 | 0.078 |
| 0.17 | ENTER | Logistic Regression | 0.027 | 0.096 |
| 0.18 | ENTER | Logistic Regression | 0.031 | 0.11 |
| 0.19 | ENTER | Logistic Regression | 0.035 | 0.123 |
| 0.2 | ENTER | Logistic Regression | 0.039 | 0.138 |
| 0.21 | ENTER | Logistic Regression | 0.041 | 0.143 |
| 0.22 | ENTER | Logistic Regression | 0.046 | 0.163 |
| 0.23 | ENTER | Logistic Regression | 0.055 | 0.195 |
| 0.24 | ENTER | Logistic Regression | 0.059 | 0.207 |
| 0.25 | ENTER | Logistic Regression | 0.066 | 0.234 |
| 0.26 | ENTER | Logistic Regression | 0.073 | 0.258 |
| 0.27 | ENTER | Logistic Regression | 0.081 | 0.285 |
| 0.28 | ENTER | Logistic Regression | 0.09 | 0.316 |
| 0.29 | ENTER | Logistic Regression | 0.088 | 0.31 |
| 0.3 | ENTER | Logistic Regression | 0.084 | 0.296 |
| 0.31 | ENTER | Logistic Regression | 0.078 | 0.276 |
| 0.32 | ENTER | Logistic Regression | 0.063 | 0.221 |
| 0.33 | ENTER | Logistic Regression | 0.06 | 0.212 |
| 0.34 | ENTER | Logistic Regression | 0.055 | 0.193 |
| 0.35 | ENTER | Logistic Regression | 0.049 | 0.174 |
| 0.36 | ENTER | Logistic Regression | 0.046 | 0.163 |
| 0.37 | ENTER | Logistic Regression | 0.048 | 0.168 |
| 0.38 | ENTER | Logistic Regression | 0.044 | 0.155 |
| 0.39 | ENTER | Logistic Regression | 0.044 | 0.156 |
| 0.4 | ENTER | Logistic Regression | 0.041 | 0.144 |
| 0.41 | ENTER | Logistic Regression | 0.033 | 0.118 |
| 0.42 | ENTER | Logistic Regression | 0.024 | 0.085 |
| 0.43 | ENTER | Logistic Regression | 0.019 | 0.067 |
| 0.44 | ENTER | Logistic Regression | 0.016 | 0.055 |
| 0.45 | ENTER | Logistic Regression | 0.01 | 0.035 |
| 0.46 | ENTER | Logistic Regression | 0.006 | 0.02 |
| 0.47 | ENTER | Logistic Regression | -0.003 | -0.009 |
| 0.48 | ENTER | Logistic Regression | -0.013 | -0.046 |
| 0.49 | ENTER | Logistic Regression | -0.014 | -0.05 |
| 0.5 | ENTER | Logistic Regression | -0.025 | -0.088 |
| 0 | ENTER | Random Forest | 0 | 0 |
| 0.01 | ENTER | Random Forest | 0 | 0 |
| 0.02 | ENTER | Random Forest | 0 | 0.001 |
| 0.03 | ENTER | Random Forest | 0.001 | 0.002 |
| 0.04 | ENTER | Random Forest | 0.001 | 0.004 |
| 0.05 | ENTER | Random Forest | 0.002 | 0.008 |
| 0.06 | ENTER | Random Forest | 0.004 | 0.013 |
| 0.07 | ENTER | Random Forest | 0.005 | 0.017 |
| 0.08 | ENTER | Random Forest | 0.004 | 0.016 |
| 0.09 | ENTER | Random Forest | 0.006 | 0.021 |
| 0.1 | ENTER | Random Forest | 0.008 | 0.03 |
| 0.11 | ENTER | Random Forest | 0.012 | 0.043 |
| 0.12 | ENTER | Random Forest | 0.015 | 0.054 |
| 0.13 | ENTER | Random Forest | 0.019 | 0.066 |
| 0.14 | ENTER | Random Forest | 0.022 | 0.079 |
| 0.15 | ENTER | Random Forest | 0.024 | 0.086 |
| 0.16 | ENTER | Random Forest | 0.029 | 0.101 |
| 0.17 | ENTER | Random Forest | 0.033 | 0.115 |
| 0.18 | ENTER | Random Forest | 0.034 | 0.121 |
| 0.19 | ENTER | Random Forest | 0.038 | 0.134 |
| 0.2 | ENTER | Random Forest | 0.044 | 0.154 |
| 0.21 | ENTER | Random Forest | 0.044 | 0.155 |
| 0.22 | ENTER | Random Forest | 0.045 | 0.159 |
| 0.23 | ENTER | Random Forest | 0.048 | 0.17 |
| 0.24 | ENTER | Random Forest | 0.054 | 0.191 |
| 0.25 | ENTER | Random Forest | 0.062 | 0.22 |
| 0.26 | ENTER | Random Forest | 0.068 | 0.242 |
| 0.27 | ENTER | Random Forest | 0.078 | 0.275 |
| 0.28 | ENTER | Random Forest | 0.082 | 0.291 |
| 0.29 | ENTER | Random Forest | 0.077 | 0.274 |
| 0.3 | ENTER | Random Forest | 0.072 | 0.253 |
| 0.31 | ENTER | Random Forest | 0.064 | 0.228 |
| 0.32 | ENTER | Random Forest | 0.057 | 0.202 |
| 0.33 | ENTER | Random Forest | 0.049 | 0.173 |
| 0.34 | ENTER | Random Forest | 0.04 | 0.14 |
| 0.35 | ENTER | Random Forest | 0.038 | 0.134 |
| 0.36 | ENTER | Random Forest | 0.035 | 0.122 |
| 0.37 | ENTER | Random Forest | 0.032 | 0.112 |
| 0.38 | ENTER | Random Forest | 0.027 | 0.096 |
| 0.39 | ENTER | Random Forest | 0.021 | 0.076 |
| 0.4 | ENTER | Random Forest | 0.014 | 0.048 |
| 0.41 | ENTER | Random Forest | 0.01 | 0.035 |
| 0.42 | ENTER | Random Forest | 0.007 | 0.024 |
| 0.43 | ENTER | Random Forest | 0.002 | 0.006 |
| 0.44 | ENTER | Random Forest | -0.001 | -0.004 |
| 0.45 | ENTER | Random Forest | -0.003 | -0.01 |
| 0.46 | ENTER | Random Forest | -0.007 | -0.025 |
| 0.47 | ENTER | Random Forest | -0.001 | -0.003 |
| 0.48 | ENTER | Random Forest | -0.002 | -0.008 |
| 0.49 | ENTER | Random Forest | -0.005 | -0.017 |
| 0.5 | ENTER | Random Forest | -0.013 | -0.045 |
| 0 | ENTER | XGBoost | 0 | 0 |
| 0.01 | ENTER | XGBoost | 0 | 0 |
| 0.02 | ENTER | XGBoost | 0 | 0.002 |
| 0.03 | ENTER | XGBoost | 0.001 | 0.005 |
| 0.04 | ENTER | XGBoost | 0.003 | 0.011 |
| 0.05 | ENTER | XGBoost | 0.002 | 0.008 |
| 0.06 | ENTER | XGBoost | 0.004 | 0.014 |
| 0.07 | ENTER | XGBoost | 0.007 | 0.024 |
| 0.08 | ENTER | XGBoost | 0.005 | 0.017 |
| 0.09 | ENTER | XGBoost | 0.009 | 0.031 |
| 0.1 | ENTER | XGBoost | 0.011 | 0.04 |
| 0.11 | ENTER | XGBoost | 0.012 | 0.043 |
| 0.12 | ENTER | XGBoost | 0.015 | 0.053 |
| 0.13 | ENTER | XGBoost | 0.013 | 0.046 |
| 0.14 | ENTER | XGBoost | 0.015 | 0.052 |
| 0.15 | ENTER | XGBoost | 0.02 | 0.069 |
| 0.16 | ENTER | XGBoost | 0.022 | 0.077 |
| 0.17 | ENTER | XGBoost | 0.026 | 0.091 |
| 0.18 | ENTER | XGBoost | 0.032 | 0.114 |
| 0.19 | ENTER | XGBoost | 0.034 | 0.121 |
| 0.2 | ENTER | XGBoost | 0.04 | 0.142 |
| 0.21 | ENTER | XGBoost | 0.047 | 0.165 |
| 0.22 | ENTER | XGBoost | 0.054 | 0.193 |
| 0.23 | ENTER | XGBoost | 0.053 | 0.187 |
| 0.24 | ENTER | XGBoost | 0.053 | 0.186 |
| 0.25 | ENTER | XGBoost | 0.062 | 0.218 |
| 0.26 | ENTER | XGBoost | 0.071 | 0.249 |
| 0.27 | ENTER | XGBoost | 0.082 | 0.29 |
| 0.28 | ENTER | XGBoost | 0.088 | 0.312 |
| 0.29 | ENTER | XGBoost | 0.086 | 0.303 |
| 0.3 | ENTER | XGBoost | 0.082 | 0.289 |
| 0.31 | ENTER | XGBoost | 0.075 | 0.264 |
| 0.32 | ENTER | XGBoost | 0.071 | 0.251 |
| 0.33 | ENTER | XGBoost | 0.065 | 0.229 |
| 0.34 | ENTER | XGBoost | 0.059 | 0.208 |
| 0.35 | ENTER | XGBoost | 0.049 | 0.173 |
| 0.36 | ENTER | XGBoost | 0.032 | 0.112 |
| 0.37 | ENTER | XGBoost | 0.031 | 0.108 |
| 0.38 | ENTER | XGBoost | 0.025 | 0.089 |
| 0.39 | ENTER | XGBoost | 0.021 | 0.075 |
| 0.4 | ENTER | XGBoost | 0.018 | 0.063 |
| 0.41 | ENTER | XGBoost | 0.014 | 0.05 |
| 0.42 | ENTER | XGBoost | 0.003 | 0.01 |
| 0.43 | ENTER | XGBoost | -0.001 | -0.003 |
| 0.44 | ENTER | XGBoost | -0.001 | -0.004 |
| 0.45 | ENTER | XGBoost | -0.005 | -0.018 |
| 0.46 | ENTER | XGBoost | -0.011 | -0.038 |
| 0.47 | ENTER | XGBoost | -0.019 | -0.067 |
| 0.48 | ENTER | XGBoost | -0.022 | -0.079 |
| 0.49 | ENTER | XGBoost | -0.029 | -0.101 |
| 0.5 | ENTER | XGBoost | -0.036 | -0.127 |

**eTable 6** Model coefficients and variable importance

| **Model** | **Feature** | **Logistic regression coefficient** | **Random forest importance** | **XGBoost gain** |
| --- | --- | --- | --- | --- |
| PQ-16 Cut-off | (Intercept) | -5.357 (-6.216- -5.080) | NA (NA - NA) | NA (NA - NA) |
| PQ-16 Cut-off | PQ-16 cut-off | 5.873 (5.618-6.675) | 0.276 (0.270 - 0.279) | NA (NA - NA) |
| ENTER | (Intercept) | -0.197 (-3.500 - 2.537) | NA (NA - NA) | NA (NA - NA) |
| ENTER | Adult life events | 0.261 (0.107 - 0.466) | 0.004 (0.001 - 0.013) | 0.039 (0.010 - 0.077) |
| ENTER | Age | -0.143 (-0.249 - -0.054) | 0.018 (0.002 - 0.058) | 0.129 (0.039 - 0.303) |
| ENTER | Black ethnicity | 0.000 (-0.307 - 2.326) | NA (NA - NA) | 0.008 (0.000 - 0.018) |
| ENTER | Childhood trauma | -0.005 (-0.032 - 0.000) | 0.001 (0.000 - 0.002) | 0.014 (0.002 - 0.029) |
| ENTER | DSST | -0.001 (-0.025 - 0.001) | 0.000 (0.000 - 0.001) | 0.012 (0.001 - 0.021) |
| ENTER | Daily tobacco use | -0.313 (-0.935 - 0.000) | -0.000 (-0.000 - 0.000) | 0.006 (0.000 - 0.027) |
| ENTER | Ethnic density | NA (NA - NA) | 0.002 (0.000 - 0.007) | NA (NA - NA) |
| ENTER | Ethnicity | NA (NA - NA) | 0.003 (0.001 - 0.010) | NA (NA - NA) |
| ENTER | Family history of psychosis | 0.134 (-0.117 - 1.589) | 0.000 (-0.000 - 0.002) | 0.002 (0.000 - 0.014) |
| ENTER | Female gender | NA (NA - NA) | NA (NA - NA) | 0.002 (0.000 - 0.010) |
| ENTER | First-generation migrant | -1.502 (-3.000 - -0.581) | 0.020 (0.002 - 0.079) | 0.107 (0.021 - 0.255) |
| ENTER | Gender | NA (NA - NA) | 0.000 (0.000 - 0.002) | NA (NA - NA) |
| ENTER | Heavy cannabis use | 0.000 (-0.194 - 2.880) | -0.000 (-0.000 - 0.000) | NA (NA - NA) |
| ENTER | High urbanicity | 1.842 (0.014 - 3.483) | 0.001 (-0.000 - 0.007) | 0.024 (0.000 - 0.061) |
| ENTER | Low ethnic density | -0.409 (-1.492 - 0.457) | NA (NA - NA) | 0.010 (0.001 - 0.051) |
| ENTER | Male gender | -0.236 (-1.094 - 0.000) | NA (NA - NA) | 0.007 (0.001 - 0.026) |
| ENTER | Medium ethnic density | 0.612 (0.000 - 3.112) | NA (NA - NA) | 0.006 (0.000 - 0.022) |
| ENTER | Mixed ethnicity | -0.276 (-1.695 - 0.196) | NA (NA - NA) | 0.003 (0.000 - 0.020) |
| ENTER | Non-binary gender | 0.182 (-0.115 - 1.113) | NA (NA - NA) | 0.002 (0.000 - 0.013) |
| ENTER | North African first-generation migrant | 0.000 (-1.450 - 3.638) | 0.000 (-0.000 - 0.000) | NA (NA - NA) |
| ENTER | North African second-generation migrant | 0.000 (0.000 - 3.782) | 0.000 (-0.000 - 0.000) | NA (NA - NA) |
| ENTER | Other ethnicity | 1.240 (0.000 - 2.977) | NA (NA - NA) | 0.017 (0.000 - 0.035) |
| ENTER | Other gender | -1.781 (-3.957 - 0.760) | NA (NA - NA) | 0.002 (0.001 - 0.008) |
| ENTER | PQ-16 distress | 0.060 (0.009 - 0.113) | 0.028 (0.015 - 0.065) | 0.159 (0.115 - 0.291) |
| ENTER | PQ-16 score | 0.084 (0.000 - 0.306) | 0.019 (0.011 - 0.077) | 0.086 (0.028 - 0.385) |
| ENTER | Parent severe mental illness | 1.142 (0.156 - 2.706) | 0.007 (0.000 - 0.069) | 0.067 (0.002 - 0.244) |
| ENTER | Paternal age >=35 years | 1.337 (0.284 - 2.343) | 0.018 (0.001 - 0.105) | 0.082 (0.017 - 0.301) |
| ENTER | Paternal age >=45 years | 0.112 (-0.139 - 2.170) | 0.000 (-0.000 - 0.009) | 0.011 (0.000 - 0.032) |
| ENTER | Paternal socioeconomic status | 0.000 (-0.369 - 0.217) | 0.000 (-0.000 - 0.000) | 0.001 (0.000 - 0.007) |
| ENTER | Pollution | -0.101 (-0.192 - -0.039) | 0.009 (0.002 - 0.025) | 0.072 (0.027 - 0.162) |
| ENTER | Second-generation migrant | 0.284 (0.000 - 1.392) | 0.002 (0.000 - 0.017) | 0.008 (0.001 - 0.056) |
| ENTER | Trait anhedonia | 0.000 (-0.017 - 0.010) | 0.000 (-0.000 - 0.001) | 0.006 (0.001 - 0.021) |
| ENTER | White ethnicity | 0.331 (0.000 - 0.993) | NA (NA - NA) | 0.007 (0.001 - 0.033) |

**eTable 7** Subgroup performance across age, country, gender and ethnicity for ENTER

| **Model** | **Subgroup** | **n** | **Event rate** | **Sparse?** | **C-index** | **Balanced accuracy** | **Brier score** | **Calibration intercept** | **Calibration slope** |
| --- | --- | --- | --- | --- | --- | --- | --- | --- | --- |
| Logistic Regression | 18 and over | 478 | 0.24 | No | 0.741 (0.690-0.788) | 72.4% (67.9%-76.8%) | 0.20 (0.18-0.22) | -1.07 (-1.37--0.78) | 0.54 (0.40-0.70) |
| Logistic Regression | Under 18 | 16 | 0.44 | Yes | 0.621 (0.444-0.844) | 61.3% (50.0%-77.3%) | 0.36 (0.20-0.53) | -1.90 (-4.43--0.11) | 0.17 (-0.81-0.98) |
| Random Forest | 18 and over | 478 | 0.24 | No | 0.739 (0.688-0.786) | 73.1% (69.0%-77.2%) | 0.18 (0.17-0.20) | -0.76 (-1.00--0.54) | 1.04 (0.76-1.35) |
| Random Forest | Under 18 | 16 | 0.44 | Yes | 0.623 (0.438-0.873) | 55.5% (50.0%-68.8%) | 0.28 (0.20-0.37) | -0.75 (-2.05-0.43) | 2.18 (-2.44-2.53) |
| XGBoost | 18 and over | 478 | 0.24 | No | 0.731 (0.680-0.780) | 70.9% (66.1%-75.4%) | 0.20 (0.18-0.22) | -0.98 (-1.27--0.73) | 0.62 (0.46-0.79) |
| XGBoost | Under 18 | 16 | 0.44 | Yes | 0.618 (0.436-0.855) | 61.2% (50.0%-75.0%) | 0.35 (0.20-0.50) | -1.44 (-3.00--0.04) | 0.71 (-1.28-1.29) |
| Logistic Regression | Female | 356 | 0.25 | No | 0.727 (0.664-0.785) | 72.8% (67.3%-78.3%) | 0.21 (0.18-0.24) | -1.08 (-1.45--0.75) | 0.47 (0.31-0.65) |
| Logistic Regression | Male | 100 | 0.18 | No | 0.771 (0.631-0.896) | 69.7% (57.6%-81.3%) | 0.16 (0.12-0.21) | -1.23 (-1.91--0.63) | 0.78 (0.35-1.44) |
| Logistic Regression | Non-binary | 30 | 0.43 | No | 0.600 (0.486-0.767) | 59.9% (45.0%-75.0%) | 0.30 (0.21-0.41) | -0.92 (-2.11-0.10) | 0.16 (-0.44-0.79) |
| Random Forest | Female | 356 | 0.25 | No | 0.727 (0.670-0.781) | 73.1% (68.4%-77.4%) | 0.19 (0.17-0.20) | -0.74 (-1.01--0.50) | 0.97 (0.69-1.30) |
| Random Forest | Male | 100 | 0.18 | No | 0.767 (0.633-0.886) | 70.1% (58.2%-80.6%) | 0.17 (0.14-0.20) | -1.11 (-1.70--0.61) | 1.24 (0.50-2.52) |
| Random Forest | Non-binary | 30 | 0.43 | No | 0.614 (0.472-0.804) | 66.5% (52.5%-80.6%) | 0.24 (0.19-0.29) | -0.30 (-1.14-0.51) | 0.73 (-0.53-2.27) |
| XGBoost | Female | 356 | 0.25 | No | 0.716 (0.657-0.772) | 70.4% (65.0%-75.6%) | 0.21 (0.19-0.23) | -1.01 (-1.33--0.69) | 0.55 (0.38-0.74) |
| XGBoost | Male | 100 | 0.18 | No | 0.763 (0.616-0.892) | 69.4% (56.7%-81.6%) | 0.17 (0.13-0.21) | -1.24 (-1.89--0.68) | 0.83 (0.32-1.53) |
| XGBoost | Non-binary | 30 | 0.43 | No | 0.604 (0.483-0.774) | 68.7% (52.3%-83.3%) | 0.28 (0.20-0.36) | -0.58 (-1.60-0.43) | 0.29 (-0.46-1.04) |
| Logistic Regression | Asian | 83 | 0.14 | No | 0.662 (0.466-0.814) | 60.3% (46.6%-75.4%) | 0.16 (0.11-0.21) | -0.92 (-1.96--0.04) | 0.49 (0.00-0.78) |
| Logistic Regression | Black | 15 | 0.33 | Yes | 0.708 (0.444-1.000) | 74.9% (47.2%-95.5%) | 0.21 (0.10-0.34) | -0.39 (-2.25-1.10) | 3.13 (0.03-40.02) |
| Logistic Regression | Mixed | 24 | 0.29 | Yes | 0.614 (0.476-0.813) | 41.9% (19.4%-65.6%) | 0.33 (0.22-0.44) | -1.08 (-2.78-0.40) | 0.02 (-0.63-0.62) |
| Logistic Regression | Other | 16 | 0.38 | Yes | 0.637 (0.467-0.896) | 58.6% (31.2%-83.3%) | 0.31 (0.17-0.48) | -0.77 (-3.71-1.06) | 0.09 (-0.90-1.58) |
| Logistic Regression | White | 355 | 0.26 | No | 0.769 (0.714-0.822) | 75.4% (70.8%-79.9%) | 0.20 (0.18-0.23) | -1.19 (-1.52--0.89) | 0.62 (0.45-0.83) |
| Random Forest | Asian | 83 | 0.14 | No | 0.697 (0.528-0.845) | 73.5% (57.4%-87.8%) | 0.14 (0.11-0.18) | -0.95 (-1.75--0.32) | 0.80 (0.14-1.69) |
| Random Forest | Black | 15 | 0.33 | Yes | 0.764 (0.500-1.000) | 80.0% (63.6%-95.0%) | 0.18 (0.10-0.26) | -0.29 (-1.64-0.81) | 29.65 (0.26-49.30) |
| Random Forest | Mixed | 24 | 0.29 | Yes | 0.600 (0.411-0.815) | 60.4% (41.7%-76.3%) | 0.27 (0.21-0.33) | -0.79 (-2.00-0.23) | 0.18 (-1.26-1.19) |
| Random Forest | Other | 16 | 0.38 | Yes | 0.647 (0.467-0.900) | 67.8% (41.7%-91.7%) | 0.24 (0.14-0.36) | 0.11 (-1.32-1.34) | 1.14 (-0.72-2.91) |
| Random Forest | White | 355 | 0.26 | No | 0.755 (0.699-0.810) | 72.1% (67.4%-76.5%) | 0.19 (0.17-0.20) | -0.80 (-1.06--0.55) | 1.22 (0.89-1.66) |
| XGBoost | Asian | 83 | 0.14 | No | 0.675 (0.474-0.818) | 66.5% (51.2%-81.7%) | 0.16 (0.11-0.21) | -0.98 (-1.87--0.17) | 0.45 (0.10-0.89) |
| XGBoost | Black | 15 | 0.33 | Yes | 0.738 (0.444-1.000) | 65.0% (37.5%-90.9%) | 0.22 (0.11-0.35) | -0.35 (-2.22-1.12) | 9.10 (-0.07-75.60) |
| XGBoost | Mixed | 24 | 0.29 | Yes | 0.601 (0.444-0.807) | 49.4% (25.0%-71.6%) | 0.31 (0.22-0.41) | -1.05 (-2.47-0.21) | -0.01 (-0.75-0.61) |
| XGBoost | Other | 16 | 0.38 | Yes | 0.634 (0.444-0.875) | 55.1% (28.6%-81.3%) | 0.29 (0.15-0.45) | -0.34 (-2.37-1.28) | 0.41 (-0.74-1.59) |
| XGBoost | White | 355 | 0.26 | No | 0.755 (0.701-0.808) | 73.3% (68.3%-77.9%) | 0.20 (0.18-0.23) | -1.07 (-1.36--0.79) | 0.73 (0.54-0.97) |
| Logistic Regression | Italy | 110 | 0.18 | No | 0.683 (0.545-0.815) | 75.6% (63.6%-86.4%) | 0.17 (0.13-0.21) | -0.93 (-1.63--0.29) | 0.47 (0.08-0.95) |
| Logistic Regression | UK | 385 | 0.27 | No | 0.743 (0.689-0.794) | 71.3% (66.2%-76.1%) | 0.21 (0.19-0.24) | -1.12 (-1.47--0.81) | 0.51 (0.37-0.68) |
| Random Forest | Italy | 110 | 0.18 | No | 0.692 (0.566-0.816) | 71.5% (59.9%-81.8%) | 0.17 (0.15-0.20) | -1.01 (-1.59--0.51) | 1.07 (0.38-2.04) |
| Random Forest | UK | 385 | 0.27 | No | 0.735 (0.682-0.786) | 72.6% (68.4%-76.8%) | 0.19 (0.17-0.21) | -0.69 (-0.94--0.45) | 0.97 (0.71-1.29) |
| XGBoost | Italy | 110 | 0.18 | No | 0.659 (0.526-0.782) | 64.4% (52.7%-76.4%) | 0.18 (0.14-0.22) | -0.93 (-1.54--0.36) | 0.49 (0.06-0.99) |
| XGBoost | UK | 385 | 0.27 | No | 0.734 (0.680-0.786) | 71.6% (66.7%-76.2%) | 0.21 (0.19-0.24) | -1.02 (-1.31--0.71) | 0.60 (0.43-0.78) |

| **eTable 8** Bootstrapped pairwise comparisons for subgroup analyses across age, country, gender and ethnicity for ENTER models. Analyses were restricted to comparisons between subgroups that were not classified as sparse. | | | | | | | | | | | | |
| --- | --- | --- | --- | --- | --- | --- | --- | --- | --- | --- | --- | --- |
| **Model** | **Subgroup A** | **Subgroup B** | **C-index** | | **Balanced accuracy** | | **Brier score** | | **Calibration intercept** | | **Calibration slope** | |
|  |  |  | **Difference (95% CIs)** | **p-value** | **Difference (95% CIs)** | **p-value** | **Difference (95% CIs)** | **p-value** | **Difference (95% CIs)** | **p-value** | **Difference (95% CIs)** | **p-value** |
| Logistic regression | Female | Male | -0.047 (-0.187-0.115) | 0.52 | 0.027 (-0.102-0.167) | 0.693 | 0.046 (-0.006-0.095) | 0.079 | 0.990 (-0.414-3.010) | 0.208 | -1.336 (-4.506-1.222) | 0.33 |
| Random forest | Female | Male | -0.040 (-0.171-0.103) | 0.556 | 0.031 (-0.083-0.153) | 0.63 | 0.021 (-0.013-0.055) | 0.227 | 0.967 (-0.848-3.896) | 0.38 | -1.206 (-6.433-2.540) | 0.62 |
| XGBoost | Female | Male | -0.045 (-0.188-0.107) | 0.55 | 0.011 (-0.123-0.150) | 0.884 | 0.042 (-0.005-0.088) | 0.079 | 0.982 (-0.437-3.003) | 0.212 | -1.285 (-4.515-1.264) | 0.368 |
| Logistic regression | Female | Non-binary | 0.161 (-0.059-0.386) | 0.155 | 0.128 (-0.032-0.291) | 0.119 | -0.094 (-0.197-0.004) | 0.066 | -1.493 (-3.641-0.863) | 0.185 | 1.777 (-1.780-5.224) | 0.296 |
| Random forest | Female | Non-binary | 0.128 (-0.077-0.346) | 0.233 | 0.063 (-0.084-0.208) | 0.405 | **-0.055 (-0.105--0.005)** | **0.033** | -1.200 (-4.032-2.096) | 0.402 | 1.362 (-5.014-6.752) | 0.615 |
| XGBoost | Female | Non-binary | 0.142 (-0.067-0.363) | 0.195 | 0.017 (-0.146-0.182) | 0.842 | -0.068 (-0.150-0.015) | 0.118 | -1.408 (-3.551-0.836) | 0.173 | 1.542 (-2.319-5.321) | 0.368 |
| Logistic regression | Male | Non-binary | 0.210 (-0.044-0.470) | 0.103 | 0.099 (-0.108-0.302) | 0.336 | **-0.140 (-0.256--0.033)** | **0.009** | -2.532 (-5.646-0.177) | 0.064 | 3.173 (-1.054-7.655) | 0.148 |
| Random forest | Male | Non-binary | 0.170 (-0.067-0.412) | 0.171 | 0.033 (-0.144-0.202) | 0.69 | **-0.076 (-0.134--0.017)** | **0.007** | -2.182 (-5.930-1.643) | 0.246 | 2.582 (-4.640-9.730) | 0.451 |
| XGBoost | Male | Non-binary | 0.185 (-0.064-0.447) | 0.14 | 0.005 (-0.196-0.204) | 0.972 | **-0.109 (-0.199--0.019)** | **0.018** | -2.387 (-5.360-0.241) | 0.071 | 2.812 (-1.593-7.434) | 0.196 |
| Logistic regression | White | Asian | 0.106 (-0.057-0.283) | 0.216 | 0.153 (-0.004-0.298) | 0.057 | 0.047 (-0.012-0.104) | 0.13 | -0.564 (-1.628-0.672) | 0.34 | 1.835 (-0.848-4.647) | 0.155 |
| Random forest | White | Asian | 0.059 (-0.095-0.228) | 0.488 | -0.013 (-0.158-0.139) | 0.851 | **0.048 (0.004-0.087)** | **0.028** | -0.543 (-2.073-1.348) | 0.476 | 1.650 (-2.330-5.533) | 0.4 |
| XGBoost | White | Asian | 0.077 (-0.076-0.232) | 0.325 | 0.067 (-0.092-0.229) | 0.42 | 0.046 (-0.009-0.100) | 0.1 | -0.471 (-1.632-0.907) | 0.417 | 1.693 (-0.936-4.514) | 0.208 |
| Logistic regression | UK | Italy | 0.059 (-0.087-0.215) | 0.449 | -0.043 (-0.156-0.086) | 0.478 | 0.044 (-0.009-0.091) | 0.087 | 0.014 (-1.025-1.256) | 0.987 | 0.266 (-2.099-2.535) | 0.794 |
| Random forest | UK | Italy | 0.047 (-0.094-0.184) | 0.507 | 0.014 (-0.101-0.137) | 0.836 | 0.019 (-0.014-0.050) | 0.253 | 0.354 (-1.201-2.438) | 0.744 | -0.140 (-4.438-3.248) | 0.998 |
| XGBoost | UK | Italy | 0.076 (-0.059-0.224) | 0.291 | 0.073 (-0.058-0.203) | 0.272 | 0.034 (-0.011-0.077) | 0.154 | -0.165 (-1.264-0.995) | 0.732 | 0.763 (-1.859-3.286) | 0.529 |

| **eTable 9** Model performance of unimodal models | | | | | | | | | | | | | | | | | | | |
| --- | --- | --- | --- | --- | --- | --- | --- | --- | --- | --- | --- | --- | --- | --- | --- | --- | --- | --- | --- |
| **Model** | **Model Type** | | **C-index** | | **Balanced Accuracy** | | **Sensitivity** | | **Specificity** | | **PPV** | | **NPV** | | **Brier Score** | | **Calibration Intercept** | | **Calibration Slope** |
| PQ-16 score | Logistic Regression | | 0.742  (0.689-0.790) | | 71.2%  (66.8%-75.6%) | | 75.0%  (67.2%-82.7%) | | 67.4%  (62.7%-72.1%) | | 43.5%  (36.9%-50.2%) | | 89.0%  (85.3%-92.6%) | | 0.22  (0.19-0.24) | | -1.39  (-1.69--1.11) | | 0.57  (0.43-0.72) |
| PQ-16 score | Random Forest | | 0.595  (0.542-0.647) | | 65.1%  (61.4%-68.7%) | | 88.7%  (82.8%-93.8%) | | 41.6%  (36.6%-46.5%) | | 33.5%  (28.5%-38.9%) | | 91.7%  (87.4%-95.6%) | | 0.24  (0.23-0.26) | | -1.20  (-1.43--0.98) | | 0.42  (0.25-0.69) |
| PQ-16 score | XGBoost | | 0.679  (0.629-0.731) | | 65.1%  (61.3%-68.9%) | | 88.6%  (82.8%-93.8%) | | 41.6%  (36.7%-46.5%) | | 33.6%  (28.6%-38.6%) | | 91.7%  (87.3%-95.6%) | | 0.22  (0.21-0.23) | | -0.98  (-1.20--0.78) | | 0.73  (0.42-1.25) |
| PQ-16 distress | Logistic Regression | | 0.759  (0.711-0.802) | | 74.4%  (70.0%-78.6%) | | 77.4%  (69.6%-84.7%) | | 71.4%  (66.7%-75.9%) | | 47.5%  (40.3%-54.5%) | | 90.5%  (86.9%-93.6%) | | 0.20  (0.18-0.23) | | -1.26  (-1.61--0.94) | | 0.48  (0.37-0.59) |
| PQ-16 distress | Random Forest | | 0.647  (0.596-0.698) | | 68.2%  (64.5%-72.0%) | | 89.4%  (83.7%-94.4%) | | 47.0%  (42.1%-52.4%) | | 35.9%  (30.5%-41.4%) | | 93.1%  (89.2%-96.4%) | | 0.22  (0.20-0.24) | | -1.11  (-1.36--0.86) | | 0.44  (0.30-0.62) |
| PQ-16 distress | XGBoost | | 0.697  (0.648-0.749) | | 68.3%  (64.5%-72.0%) | | 89.5%  (83.8%-94.6%) | | 47.1%  (42.0%-52.4%) | | 36.0%  (30.6%-41.2%) | | 93.1%  (89.4%-96.4%) | | 0.20  (0.18-0.22) | | -0.86  (-1.08--0.63) | | 0.67  (0.47-0.93) |
| DSST | Logistic Regression | | 0.538  (0.497-0.596) | | 59.1%  (54.2%-64.3%) | | 62.9%  (54.3%-71.6%) | | 55.4%  (50.4%-60.4%) | | 32.0%  (26.1%-38.3%) | | 81.7%  (76.9%-86.5%) | | 0.26  (0.24-0.29) | | -0.87  (-1.20--0.55) | | 0.09  (-0.05-0.22) |
| DSST | Random Forest | | 0.521  (0.479-0.572) | | 59.3%  (54.5%-64.3%) | | 65.4%  (56.9%-73.5%) | | 53.2%  (48.2%-58.1%) | | 31.7%  (26.1%-37.3%) | | 82.3%  (77.4%-87.0%) | | 0.30  (0.27-0.33) | | -1.02  (-1.34--0.71) | | 0.05  (-0.05-0.16) |
| DSST | XGBoost | | 0.509  (0.457-0.568) | | 56.0%  (51.0%-61.0%) | | 59.8%  (50.8%-68.1%) | | 52.1%  (46.9%-57.0%) | | 29.4%  (23.8%-34.9%) | | 79.6%  (74.5%-84.4%) | | 0.27  (0.25-0.30) | | -0.87  (-1.14--0.60) | | 0.08  (-0.06-0.22) |
| PPS | Logistic Regression | | 0.687  (0.637-0.735) | | 70.7%  (66.3%-74.9%) | | 79.8%  (72.4%-86.3%) | | 61.5%  (56.6%-66.4%) | | 40.9%  (34.8%-47.2%) | | 90.1%  (86.4%-93.5%) | | 0.20  (0.18-0.23) | | -0.63  (-0.94--0.35) | | 0.40  (0.28-0.55) |
| PPS | Random Forest | | 0.669  (0.616-0.724) | | 67.4%  (62.8%-72.1%) | | 71.7%  (63.3%-79.2%) | | 63.2%  (58.3%-68.3%) | | 39.5%  (33.2%-45.8%) | | 86.9%  (82.9%-90.8%) | | 0.18  (0.16-0.20) | | -0.24  (-0.45--0.03) | | 0.79  (0.51-1.12) |
| PPS | XGBoost | | 0.677  (0.620-0.731) | | 67.2%  (62.3%-71.8%) | | 69.4%  (61.3%-77.7%) | | 65.0%  (59.9%-69.8%) | | 39.8%  (33.5%-46.4%) | | 86.4%  (82.1%-90.4%) | | 0.19  (0.17-0.22) | | -0.51  (-0.78--0.26) | | 0.49  (0.32-0.68) |

| **eTable 10** Bootstrapped pairwise comparisons for unimodal models and ENTER models | | | | | | | | | | | |
| --- | --- | --- | --- | --- | --- | --- | --- | --- | --- | --- | --- |
| **Model A** | **Model B** | **C-index** | | **Balanced accuracy** | | **Brier score** | | **Calibration intercept** | | **Calibration slope** | |
|  |  | **Difference (95% CIs)** | **p-value** | **Difference (95% CIs)** | **p-value** | **Difference (95% CIs)** | **p-value** | **Difference (95% CIs)** | **p-value** | **Difference (95% CIs)** | **p-value** |
| PQ-16 score – logistic regression | ENTER – logistic regression | -0.001 (-0.053-0.052) | 0.945 | -0.019 (-0.072-0.037) | 0.497 | 0.016 (-0.007-0.038) | 0.163 | -0.273 (-0.766-0.221) | 0.263 | 0.170 (-0.713-1.035) | 0.72 |
| PQ-16 score – logistic regression | ENTER – random forest | 0.001 (-0.041-0.042) | 0.943 | 0.008 (-0.041-0.055) | 0.742 | **0.032 (0.014-0.050)** | **<0.001** | 0.345 (-0.127-0.830) | 0.149 | **-1.456 (-2.434--0.569)** | **0.002** |
| PQ-16 score – logistic regression | ENTER – XGBoost | 0.004 (-0.047-0.054) | 0.85 | 0.001 (-0.054-0.055) | 0.941 | 0.017 (-0.004-0.039) | 0.114 | -0.215 (-0.728-0.269) | 0.405 | 0.016 (-0.857-0.909) | 0.967 |
| PQ-16 score – random forest | ENTER – logistic regression | **-0.148 (-0.209--0.088)** | **<0.001** | **-0.091 (-0.139--0.042)** | **<0.001** | **0.041 (0.019-0.063)** | **0.001** | -0.254 (-0.954-0.344) | 0.455 | 0.131 (-0.969-1.357) | 0.866 |
| PQ-16 score – random forest | ENTER – random forest | **-0.149 (-0.208--0.093)** | **<0.001** | **-0.066 (-0.109--0.022)** | **0.004** | **0.058 (0.042-0.076)** | **<0.001** | 0.396 (-0.246-1.045) | 0.207 | **-1.552 (-2.766--0.313)** | **0.013** |
| PQ-16 score – random forest | ENTER – XGBoost | **-0.143 (-0.201--0.084)** | **<0.001** | **-0.072 (-0.118--0.026)** | **0.003** | **0.043 (0.023-0.063)** | **0.001** | -0.192 (-0.843-0.365) | 0.55 | -0.029 (-1.065-1.133) | 0.922 |
| PQ-16 score – XGBoost | ENTER – logistic regression | **-0.058 (-0.113--0.006)** | **0.028** | **-0.093 (-0.141--0.046)** | **<0.001** | 0.019 (-0.001-0.040) | 0.072 | **-0.846 (-2.128--0.013)** | **0.046** | 1.555 (-0.076-3.962) | 0.07 |
| PQ-16 score – XGBoost | ENTER – random forest | **-0.058 (-0.106--0.010)** | **0.019** | **-0.066 (-0.109--0.021)** | **0.005** | **0.036 (0.021-0.049)** | **<0.001** | -0.229 (-1.546-0.608) | 0.734 | -0.066 (-1.693-2.337) | 0.853 |
| PQ-16 score – XGBoost | ENTER – XGBoost | **-0.052 (-0.102-0.003)** | **0.068** | **-0.071 (-0.118--0.025)** | **0.005** | **0.020 (0.002-0.038)** | **0.036** | -0.825 (-2.110-0.032) | 0.06 | 1.473 (-0.192-3.873) | 0.092 |
| PQ-16 distress – logistic regression | ENTER – logistic regression | 0.016 (-0.033-0.066) | 0.526 | -0.010 (-0.062-0.043) | 0.724 | 0.002 (-0.021-0.025) | 0.894 | -0.048 (-0.493-0.406) | 0.842 | -0.032 (-0.850-0.768) | 0.932 |
| PQ-16 distress – logistic regression | ENTER – random forest | 0.016 (-0.028-0.058) | 0.43 | 0.015 (-0.034-0.063) | 0.563 | **0.018 (0.000-0.037)** | **0.045** | **0.595 (0.137-1.102)** | **0.01** | **-1.701 (-2.728--0.757)** | **<0.001** |
| PQ-16 distress – logistic regression | ENTER – XGBoost | 0.023 (-0.026-0.071) | 0.351 | 0.010 (-0.045-0.062) | 0.729 | 0.002 (-0.018-0.025) | 0.825 | 0.003 (-0.448-0.443) | 0.96 | -0.172 (-1.003-0.648) | 0.674 |
| PQ-16 distress – random forest | ENTER – logistic regression | **-0.100 (-0.161--0.041)** | **<0.001** | **-0.061 (-0.109--0.013)** | **0.013** | 0.022 (-0.001-0.045) | 0.065 | -0.162 (-0.786-0.379) | 0.596 | 0.104 (-0.877-1.194) | 0.872 |
| PQ-16 distress – random forest | ENTER – random forest | **-0.099 (-0.157--0.040)** | **0.001** | -0.034 (-0.079-0.011) | 0.144 | **0.038 (0.019-0.057)** | **<0.001** | 0.463 (-0.185-1.087) | 0.155 | **-1.534 (-2.735--0.292)** | **0.013** |
| PQ-16 distress – random forest | ENTER – XGBoost | **-0.094 (-0.154--0.031)** | **0.005** | -0.041 (-0.089-0.010) | 0.119 | **0.023 (0.001-0.045)** | **0.043** | -0.102 (-0.735-0.460) | 0.765 | -0.052 (-1.109-1.034) | 0.919 |
| PQ-16 distress – XGBoost | ENTER – logistic regression | -0.043 (-0.098-0.011) | 0.12 | **-0.061 (-0.108--0.013)** | **0.013** | 0.001 (-0.021-0.022) | 0.965 | -0.420 (-1.172-0.202) | 0.21 | 0.898 (-0.309-2.273) | 0.163 |
| PQ-16 distress – XGBoost | ENTER – random forest | -0.042 (-0.093-0.010) | 0.121 | -0.034 (-0.079-0.011) | 0.123 | **0.017 (0.001-0.032)** | **0.033** | 0.212 (-0.612-0.905) | 0.528 | -0.748 (-2.119-0.787) | 0.274 |
| PQ-16 distress – XGBoost | ENTER – XGBoost | -0.038 (-0.093-0.014) | 0.164 | -0.041 (-0.091-0.007) | 0.09 | 0.002 (-0.018-0.022) | 0.869 | -0.357 (-1.051-0.289) | 0.293 | 0.738 (-0.485-2.047) | 0.255 |
| DSST – random forest | ENTER – logistic regression | **-0.225 (-0.293--0.154)** | **<0.001** | **-0.153 (-0.215--0.091)** | **<0.001** | **0.097 (0.066-0.130)** | **<0.001** | **1.288 (0.849-1.755)** | **<0.001** | **-2.752 (-3.721--1.838)** | **<0.001** |
| DSST – random forest | ENTER – random forest | **-0.225 (-0.298--0.150)** | **<0.001** | **-0.126 (-0.190--0.059)** | **<0.001** | **0.114 (0.083-0.144)** | **<0.001** | **1.929 (1.324-2.567)** | **<0.001** | **-4.420 (-5.769--3.112)** | **<0.001** |
| DSST – random forest | ENTER – XGBoost | **-0.219 (-0.289--0.144)** | **<0.001** | **-0.131 (-0.199--0.066)** | **<0.001** | **0.098 (0.066-0.130)** | **<0.001** | **1.343 (0.872-1.818)** | **<0.001** | **-2.900 (-3.875--1.919)** | **<0.001** |
| DSST – XGBoost | ENTER – logistic regression | **-0.254 (-0.329--0.178)** | **<0.001** | **-0.173 (-0.239--0.106)** | **<0.001** | **0.071 (0.040-0.102)** | **<0.001** | **1.262 (0.799-1.770)** | **<0.001** | **-2.683 (-3.731--1.654)** | **<0.001** |
| DSST – XGBoost | ENTER – random forest | **-0.254 (-0.328--0.179)** | **<0.001** | **-0.148 (-0.210--0.085)** | **<0.001** | **0.088 (0.062-0.114)** | **<0.001** | **1.902 (1.298-2.598)** | **<0.001** | **-4.344 (-5.799--3.034)** | **<0.001** |
| DSST – XGBoost | ENTER – XGBoost | **-0.247 (-0.321--0.176)** | **<0.001** | **-0.153 (-0.217--0.091)** | **<0.001** | **0.072 (0.043-0.101)** | **<0.001** | **1.309 (0.838-1.825)** | **<0.001** | **-2.812 (-3.902--1.781)** | **<0.001** |
| PPS – logistic regression | ENTER – logistic regression | **-0.052 (-0.081--0.025)** | **<0.001** | -0.020 (-0.059-0.017) | 0.292 | -0.001 (-0.013-0.012) | 0.904 | **0.516 (0.299-0.736)** | **<0.001** | **-0.685 (-1.099--0.264)** | **0.001** |
| PPS – logistic regression | ENTER – random forest | **-0.047 (-0.086--0.010)** | **0.01** | -0.023 (-0.065-0.017) | 0.28 | **0.018 (0.004-0.033)** | **0.012** | **1.135 (0.740-1.566)** | **<0.001** | **-2.348 (-3.213--1.546)** | **<0.001** |
| PPS – logistic regression | ENTER – XGBoost | **-0.041 (-0.070--0.010)** | **0.006** | -0.005 (-0.046-0.037) | 0.815 | -0.001 (-0.014-0.013) | 0.942 | **0.533 (0.299-0.788)** | **<0.001** | **-0.763 (-1.244--0.283)** | **<0.001** |
| PPS – random forest | ENTER – logistic regression | **-0.071 (-0.106--0.035)** | **<0.001** | **-0.052 (-0.097--0.007)** | **0.022** | **-0.025 (-0.044--0.005)** | **0.012** | 0.091 (-0.258-0.415) | 0.581 | **1.156 (0.188-2.290)** | **0.019** |
| PPS – random forest | ENTER – random forest | **-0.066 (-0.104--0.029)** | **<0.001** | -0.056 (-0.103--0.007) | 0.028 | -0.007 (-0.019-0.006) | 0.3 | **0.710 (0.334-1.133)** | **<0.001** | -0.508 (-1.500-0.518) | 0.334 |
| PPS – random forest | ENTER – XGBoost | **-0.060 (-0.095--0.028)** | **<0.001** | -0.038 (-0.084-0.008) | 0.107 | **-0.025 (-0.043--0.008)** | **0.006** | 0.113 (-0.209-0.420) | 0.469 | **1.056 (0.094-2.071)** | **0.029** |
| PPS – XGBoost | ENTER – logistic regression | **-0.063 (-0.094--0.032)** | **<0.001** | **-0.056 (-0.097--0.016)** | **0.008** | -0.010 (-0.024-0.005) | 0.21 | **0.457 (0.224-0.711)** | **<0.001** | -0.410 (-0.938-0.115) | 0.125 |
| PPS – XGBoost | ENTER – random forest | **-0.058 (-0.093--0.021)** | **0.001** | **-0.058 (-0.104--0.010)** | **0.013** | 0.009 (-0.005-0.022) | 0.183 | **1.070 (0.688-1.488)** | **<0.001** | **-2.063 (-2.892--1.245)** | **<0.001** |
| PPS – XGBoost | ENTER – XGBoost | **-0.052 (-0.084--0.022)** | **0.002** | -0.041 (-0.087-0.004) | 0.084 | -0.010 (-0.023-0.003) | 0.161 | **0.475 (0.243-0.731)** | **<0.001** | -0.494 (-1.023-0.011) | 0.054 |

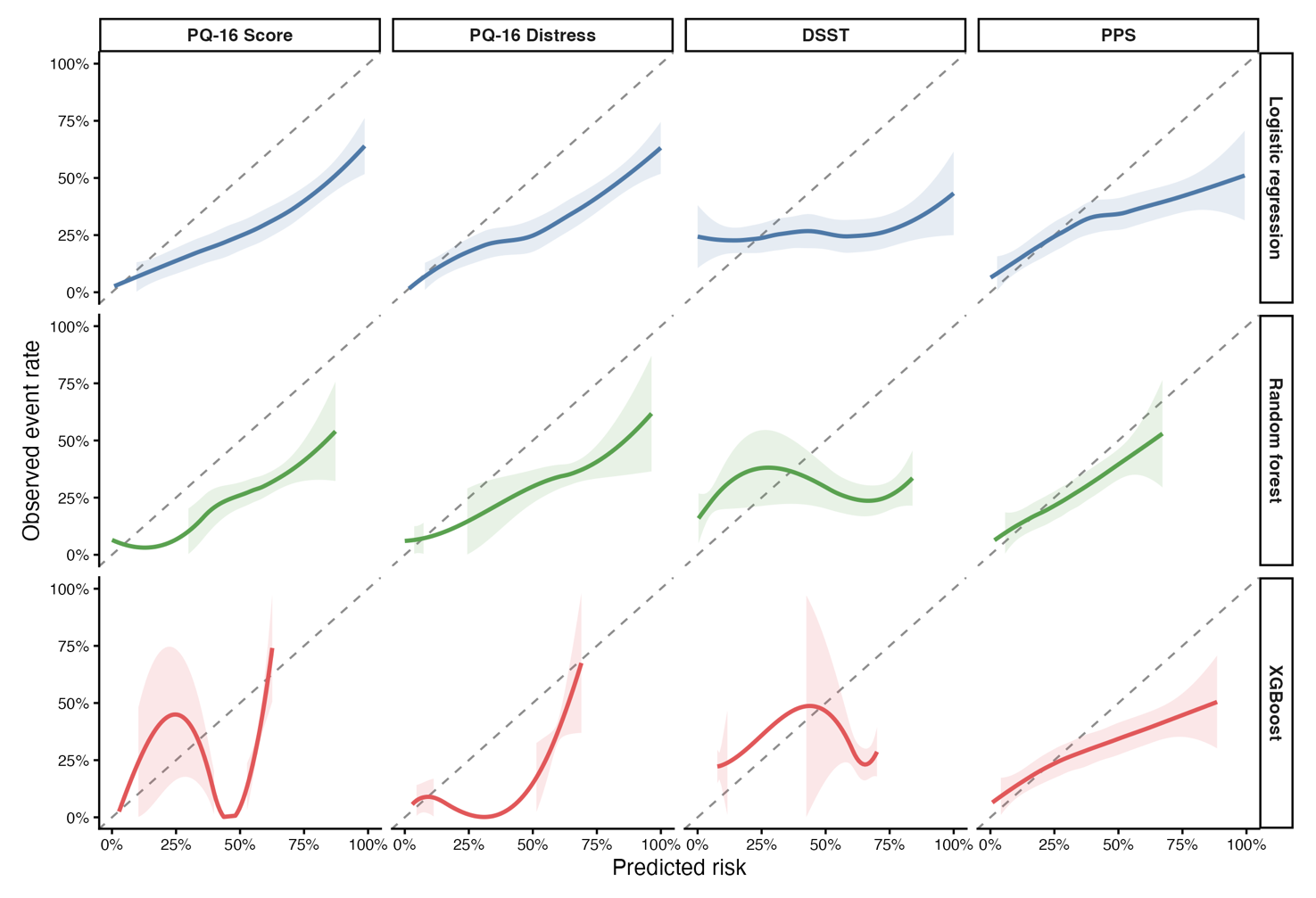

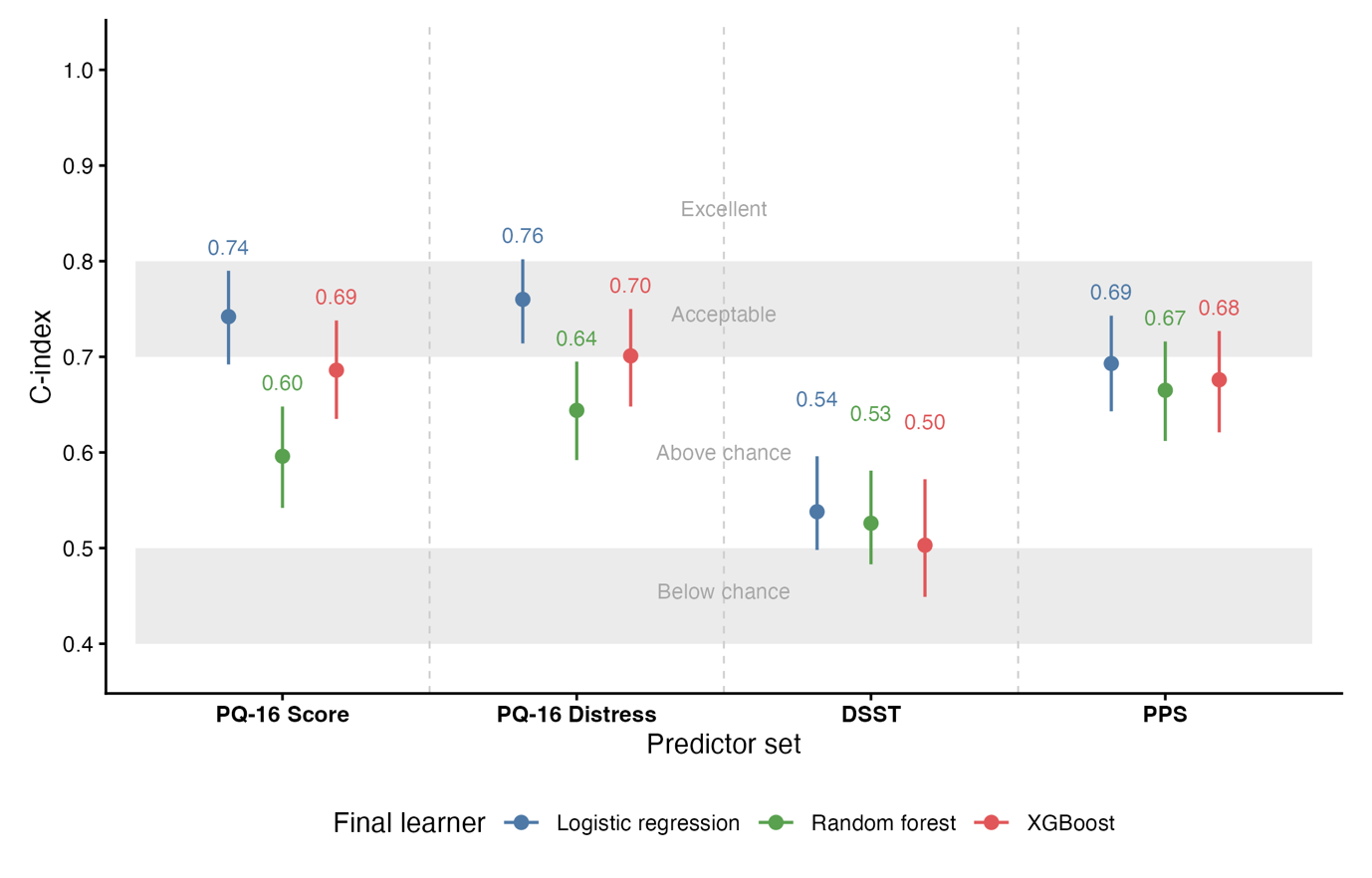
**eFigure 2** Performance summary of unimodal models. **A** Discrimination performance. **B** Calibration plots. Loess curves using pooled predictions and observed outcomes from held-out test data across cross-validation folds. Perfect calibration falls along the dashed diagonal line.

**A**

**B**
